# Contact structure and population immunity shape the selective advantage of emerging variants

**DOI:** 10.1101/2025.11.07.25339691

**Authors:** Pourya Toranj Simin, Juliana C. Taube, Elisabeta Vergu, Lulla Opatowski, Shweta Bansal, Chiara Poletto

**Affiliations:** Sorbonne Université, INSERM, Institut Pierre Louis d’Epidémiologie et de Santé Publique, Paris, France; Department of Biology, Georgetown University, Washington, DC, United States; Université Paris-Saclay, INRAE, MaIAGE, 78350 Jouy-en-Josas, France; Epidemiology and Modelling of Antibiotic Evasion Unit, Institute Pasteur, 75475 Paris Cedex 15, France; CESP, UMR1018, Université de Versailles Saint Quentin, Inserm, Paris Saclay; Department of Molecular Medicine, University of Padova, 35121 Padova, Italy

## Abstract

Epidemics are shaped by the interplay between host and pathogen population characteristics, which are themselves intertwined. In particular, host behavior and immunity profile shape pathogen population structure by affecting both the likelihood of new variant emergence and its subsequent dynamics. Theoretical studies provide a fragmented description of this complex dynamical dependency, and the empirical evidence is limited. The SARS-CoV-2 pandemic presents an unprecedented opportunity to study variant emergence. The relative growth of emerging variants over resident ones, i.e., the selection coefficient, showed spatial variations that could be associated with population immunity and the mean and dispersion of contacts, which varied greatly according to epidemic intensity and human response. We first investigated the impact of these three features on the selection coefficient in a stochastic network-based model of new variant emergence, which incorporates tunable mean and dispersion of contacts. Results systematically chart the parameter space, characterise transitions between previously identified regimes, and quantify their strength. Mean contact was positively associated with the selection coefficient in the absence of immune evasion, while the trend became non-monotonic at high immune evasion. The impact of immunity diminished as immunity increased. Importantly, greater contact dispersion slowed down the spread of variants lacking immune evasion, but this effect quickly reversed once immune evasion became non-zero. We then analysed SARS-CoV-2 Alpha emergence in the United States at the state level, examining the association of the selection coefficient with the three population features under study, reconstructed from serological, vaccination, and contact survey data. Regression analyses yielded a significant effect of population characteristics, stronger for contact structure than for immunity. Comparison with model predictions showed overall consistency. These results suggest an association between population characteristics and variant dynamics and help interpret the heterogeneity observed in variant emergence.

**Author Summary:** The response to the SARS-CoV-2 pandemic was challenged by the emergence of new phenotypic variants, which rose to dominance, altering epidemic trajectories. Quantifying the selection coefficient, i.e., the growth of the emerging variant relative to the resident one, was essential for planning interventions. Still, the same variant exhibited different selection coefficients in different populations. Strong variations in immunity and contact structure, due to the different mitigation policies, raise the hypothesis that these features may have played a role in the variant’s relative growth, as predicted in theoretical evolutionary epidemiology. Here, we focus on population immunity and mean and dispersion of contacts and consider variants with changes in transmission and/or immune evasion. Through extensive computer simulations, we quantified the association between population characteristics and the selection coefficient over a wide parameter space, reconciling previous theoretical findings. We then analysed state-level SARS-CoV-2 Alpha emergence in the United States from virological, serological, vaccination, and contact survey data. Statistical analyses were consistent with simulations and suggested that the selection coefficient was affected by contact structure more than immunity. These results provide insight that may help the design of interventions and show the importance of behavioural surveillance systems that capture evolving human contacts.

## Introduction

Epidemics are shaped by the population characteristics of both hosts and pathogens. Host characteristics, such as population immunity and contact networks at different scales, drive population-level epidemic propagation. For instance, both a large mean and a large dispersion of contacts favour widespread epidemics [1–5]. At the same time, pathogen populations often comprise multiple co-circulating variants with diverse epidemiological traits, including transmissibility, virulence, time course of infectivity, and antigenicity [6–9]. These differences influence both short-term dynamics and long-term evolutionary trajectories. Host and pathogen population characteristics are thus essential factors to account for in epidemic response, from the short-term planning of social restrictions to the long-term design of vaccines and antimicrobials [10–12].

Still, the effects of host and pathogen characteristics on epidemic dynamics are intertwined because hosts and pathogens form a coevolving system where hosts alter the conditions under which new variant traits emerge [8,13–22]. Decades of work in population genetics and invasion theory have mathematically linked population characteristics to the spread of beneficial alleles [23–27]. Leveraging this literature, theoretical evolutionary epidemiology addressed the association between the selective advantage of variant traits, such as change in transmissibility and immune evasion, and host characteristics [8,13–22]. The advantage conferred by increased transmissibility is greater at the onset of the epidemic, when a larger fraction of the population is susceptible, and transmission is higher [28]. However, for a variant with immune evasion that can reinfect previously infected individuals, the advantage increases as population immunity builds [19]. The host contact network was also found to alter trait advantage [8,14–17,20,21,29]. Broadly, the role of network structure on evolutionary dynamics has been studied for a range of processes, including Moran [30], birth-death [31,32], and epidemic processes [8,14–17,20,21,29]. In all cases, the network was found to act as either a suppressor or an amplifier of selection depending on its structure. For epidemic processes, a higher contact mean was found to increase the advantage of both more transmissible and immune-evading variants [22,33]. Contact dispersion, instead, hampered a more transmissible variant when competition is perfect [8,15] while favouring it when competition is partial (e.g., immune evasion) [20,21].

Taken together, these findings highlight a complex interplay of host and pathogen factors, yet how the results of different studies relate to one another remains unclear. A key challenge is that studies differ in both modelling approaches and metrics used to quantify variant advantage. For example, the works in refs. [8,15], about contact dispersion in the case of perfect competition, rely on the SIS model where competition arises from mutual exclusion.

In contrast, studies in refs. [20,21], on the beneficial role of contact dispersion under partial competition, use the SIR model with cross-immunity. These differences hinder reconciling the opposing results and identifying the boundaries between them. Furthermore, disentangling the relative roles of contact network and population immunity is not straightforward, as many studies addressing population immunity do not account for the contact network [13,19].

Beyond theoretical limitations, confronting these theoretical findings with empirical data remains generally difficult due to the limited availability of extensive, high-resolution microbiological records, despite some successful examples [8,28,33–35]. The emergence and spread of SARS-CoV-2 provide an unprecedented case study for examining the interplay between pathogen diversity and human population characteristics on outbreak dynamics. New, more fit variants (e.g., Alpha, Beta, Gamma, Delta, and Omicron) have repeatedly emerged, triggering a rise in cases, hospitalisations, and deaths, and prompting many countries to reinstate restrictions [7,36,37]. Virological and epidemiological investigations have shown that these variants were more transmissible than the resident virus and able to evade immunity to different degrees – limited for Alpha, more substantial for Beta, Gamma, and Omicron [36,37]. Unprecedented sequencing efforts have enabled monitoring variant dynamics across space [38,39]. The frequency of an emerging variant – the fraction of variant cases among total cases – and the selection coefficient – its growth rate relative to the resident variant – have been widely used as indicators for epidemic anticipation and control planning [7,40]. However, the same variant has exhibited different selection coefficients across geographic regions. The emergence of Alpha across US states provides a paradigmatic example. The Alpha variant was independently introduced in several US states between November and December 2020 [41–43]. In the majority of states, the local growth began in late December 2020 and took around 90 days. Fig. 1a shows the Alpha frequency for each US state [44]. Fig. 1b highlights that the selection coefficient varied from one state to another. Its value in New York, corresponding to the 10% percentile, was as low as 0.033 days^-1^, while the 90% percentile was 0.063 days^-1^ and was registered in Arizona (see Methods for the mathematical definition of the selection coefficient and the methodology used to compute it from data).

**Fig 1.**
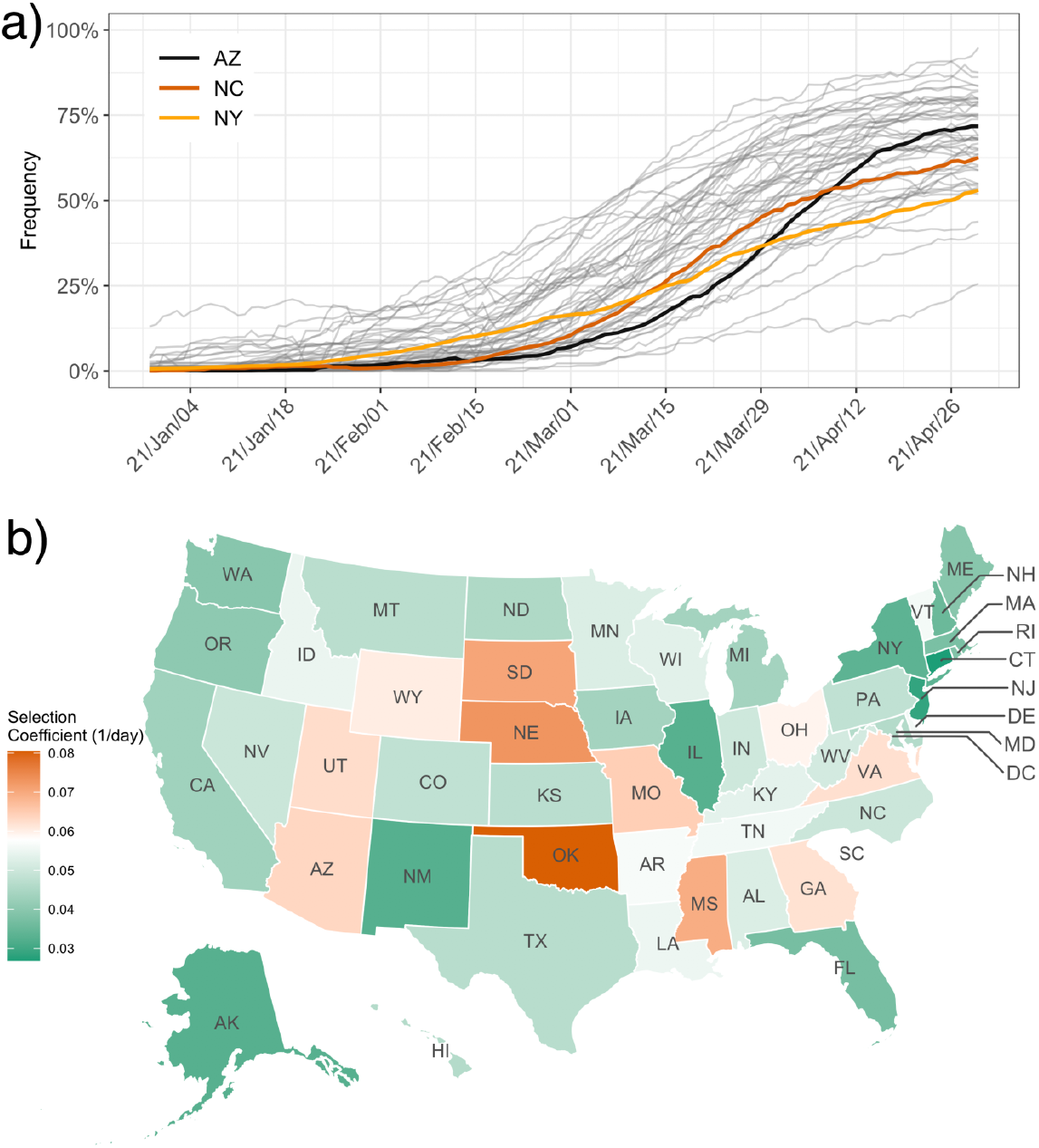
Alpha variant frequency and selection coefficients across U.S. states. a) Time series of the frequency of Alpha for each of the 50 US states, extracted from CoV-Spectrum [44]. The frequency of Alpha is defined as the incidence of Alpha divided by the total incidence. Time series are smoothed with a 21-day sliding window. Thick lines indicate the frequency in the three states where the selection coefficient is 10% percentile (New York, orange line), median (North Carolina, red line), and 90% percentile (Arizona, black line). b) Map showing the selection coefficient by state, where white color corresponds to the national mean. The selection coefficient measures the growth of the emerging variant frequency relative to the resident variant one (more details are provided in the Methods). The fitting method to compute the selection coefficient from the data is provided in the Methods section. Plots of the time series of the raw frequency for each state, together with the fit for the selection coefficient computation, are reported in Figure S1 in the Supporting Information. Base maps are generated using the R package usmap [45], based on geographic boundary data from the U.S. Census Bureau.

Similar to the US example above, other studies have highlighted spatial heterogeneities in local variant emergence dynamics [33,40,46,47]. This could be the result of variable human population characteristics. However, this link was investigated only by a few works [33,46,47]. Here, we focused on three local population characteristics – population immunity, contact mean, and its dispersion – and two variant traits – change in transmissibility and immune evasion. We analysed their association with the selection coefficient within a consistent modelling framework, which was general enough to capture the main features of acute respiratory infections (e.g., influenza, COVID-19) and disentangle the role of each factor. We then combined extensive network-based model simulations with the analysis of empirical data for Alpha emergence in the US to shed light on the source of variability observed in the data.

## Results

### Modelling variant emergence and growth

To describe variant emergence dynamics, we used the selection coefficient as a consistent metric throughout the study. The frequency of a variant is computed by its incidence divided by total incidence, i.e. 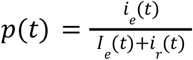, where *i*_*e*_ (*t*) and *i*_*r*_ (*t*) are the incidences of the emerging and the resident variant, respectively. The selection coefficient, *s*(*t*), is then defined as the growth of the emerging variant frequency relative to the resident variant one: 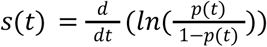 (see Methods section for details). Here, we assumed the frequency follows a logistic growth model and the selection coefficient to be constant. A positive selection coefficient indicates that the emerging variant spreads more efficiently than the resident one. The logistic growth model is widely used for its simplicity and because it synthesizes the emerging variant advantage over the resident one during a given time window [7].

We first provided theoretical predictions on the selection coefficient when the variant spreads on a local human population modelled as a contact network. We implemented a stochastic individual-based model to simulate the emergence of a variant and its co-circulation with the resident one (see Methods section for model details). As a compromise between simplicity and realism, we grouped transmission settings into two: within and outside the household (Fig. 2a). Household contacts play a central role in infection propagation due to their high clustering, proximity, and duration. On the other hand, non-household contacts are the most affected by interventions and adaptive behavioural response. Households were modelled as cliques of variable size. The number of non-household contacts (i.e. the degree of the non-household layer) was distributed as a negative binomial distribution, with mean (MD) and standard deviation (SD) explored during the analysis. On top of the network, we simulated the co-circulation of two variants interacting with cross-immunity (Fig. 2b), i.e., the immunity acquired upon infection with one variant conferred full protection to the same variant and a certain degree of protection to the other variant. This was governed by the immune evasion, or susceptibility rescale parameter, 0 ≤ σ ≤ 1. The case σ = 0 corresponds to no immune evasion (i.e., full cross immunity), while σ > 0 corresponds to some degree of immune evasion. The transmission potential of the resident variant is ruled by R_0_ which depends on transmission and recovery parameters, as well as MD and SD. By varying contact structure we explored the R_0_ value in the range [0.98 - 4.82]. The basic reproductive ratio of the emerging variant was *r*_β_ *R*_0_, where the rescaling parameter *r*_β_ quantifies the change in transmission of the emerging variant. The emerging variant was introduced by infecting 50 random individuals at a time when a fraction IM of the population had already acquired natural immunity to the resident variant. The co-circulation dynamics that followed are depicted in Fig. 2c-e for a specific set of parameters taken as an example – panel c shows the number of cases for each variant, panel d the frequency of the emerging variant, and panel e the distribution of the selection coefficient computed from each stochastic run. Other examples of the dynamics are reported in Fig. S2 in the SI.

**Fig 2.**
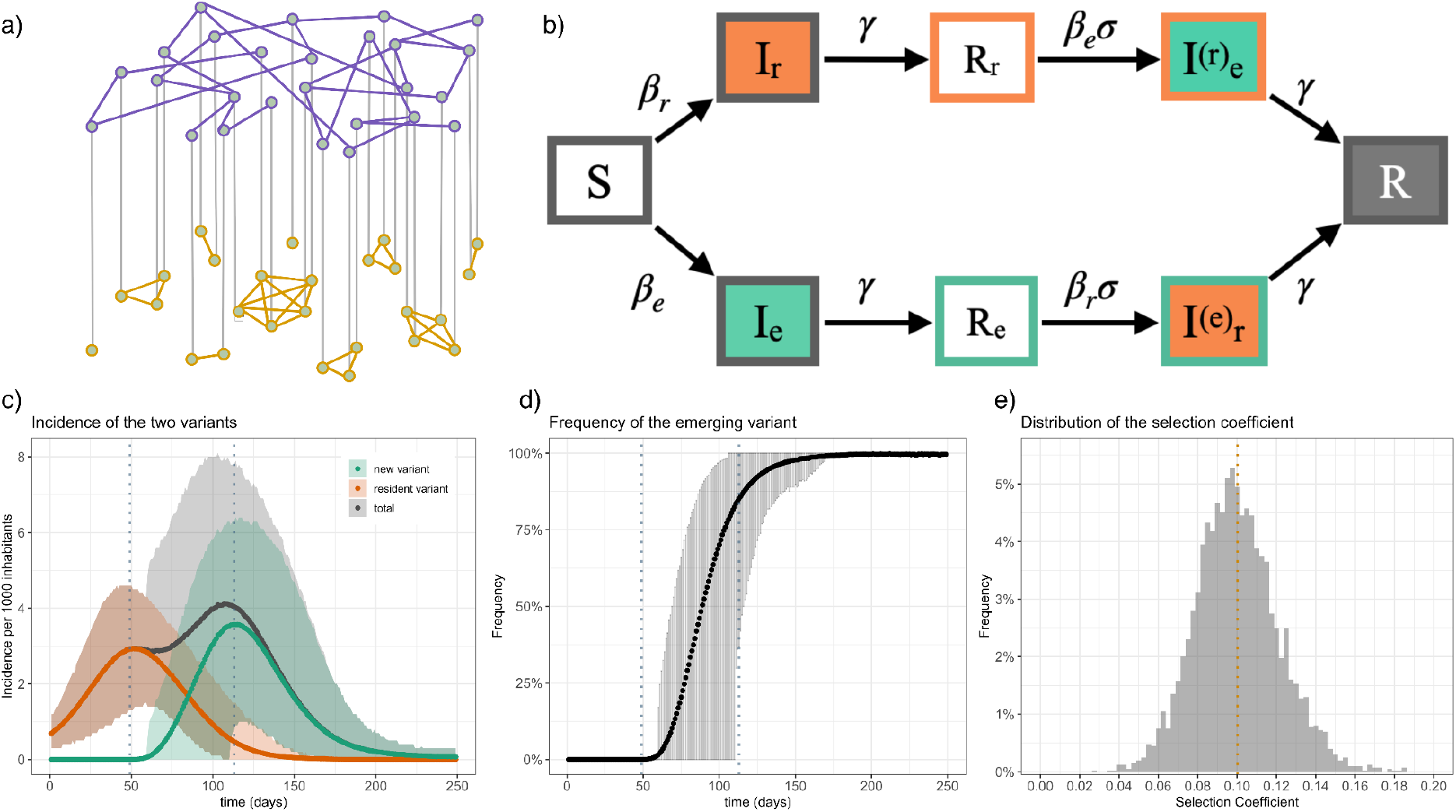
Schematic representation of the model and of the simulation output. a) Modelled contact network. This consists of two layers: the household layer (yellow links) and the social layer (purple links). b) Scheme of the 2-strain SIR model. Compartments represent: *S* (susceptible), *I*_*r*_ (Infected with resident variant), *I*_e_ (infected with emerging variant), *R*_*r*_ (recovered from resident variant), *R*_*e*_ (recovered from emerging variant), *I*^(*r*)^_*e*_(infected with emerging variant after recovering from resident variant), *I*^(*e*)^*_r_* (infected with resident variant after recovering from emerging variant), *R* (recovered from both variants). Mathematical symbols reported near the arrows indicate the transition rate: β and β indicates the transmissibility of the resident and emerging variant, respectively (β_*e*_ = *r*_β_ β_*r*_), *γ* the recovery rate (assumed to be the same for each variant), and σ the reduction in susceptibility due to cross-immunity. c) The incidence of the resident variant, the emerging variant, and the total incidence as a function of time. d) The frequency of the emerging variant, defined as the proportion of the incidence of the new variant to the total incidence. The vertical dashed lines represent the time window used to compute the selection coefficient. The shaded area shows the 95% confidence interval. e) The distribution of the computed selection coefficient of each stochastic realisation. The vertical orange line represents the mean value of the distribution, which is equal to 0.100 (1/day). Parameters used in c, d, and e are: mean number of contacts (MD = 6), standard deviation of contacts (SD = 6), immunity level (IM = 15%), immune evasion (σ = 0), transmissibility rescaling (*r*_β_ = 2), transmissibility of the resident variant β = 0. 0175, recovery rate *γ* = 0. 143 (1/7) *day*s ^−1^, and size of the network *N* = 10^4^. Statistics are computed over 100 stochastic epidemic realisations for each of the 40 stochastic generations of the network with fixed parameters, for a total of 4000 runs.

### The complex landscape of variant emergence dynamics

We used the simulation setup described above to systematically quantify the selection coefficient as we varied the levels of population immunity (IM), mean (MD), and standard deviation of contacts (SD), considering different levels of immune evasion and change in transmission. In Fig. 3a-c, we show the selection coefficient when IM, MD, and SD are varied, respectively, comparing different values of immune evasion and with the emerging variant having twice the transmissibility of the resident one. Given the strong increase in transmission, the selection coefficient was always positive. The trend of the selection coefficient in varying immunity depended on immune evasion (Fig. 3a). For low immune evasion, the selection coefficient decreased with increasing immunity, whereas for immune evasion above a certain threshold, it increased. In both cases, the selection coefficient varied more substantially when immunity was low, while it tended to plateau for high immunity. The population immunity thus reduced the advantage of a more transmissible variant (r_β_>1) if it was not able to efficiently reinfect immunised individuals, as previously shown by mean-field models and experimental observations [22,28]. In contrast, as expected, an immune-evading variant became increasingly advantaged over the resident one as population immunity increased, since this expanded the pool of individuals who can be infected by the former but not by the latter [19].

**Fig 3.**
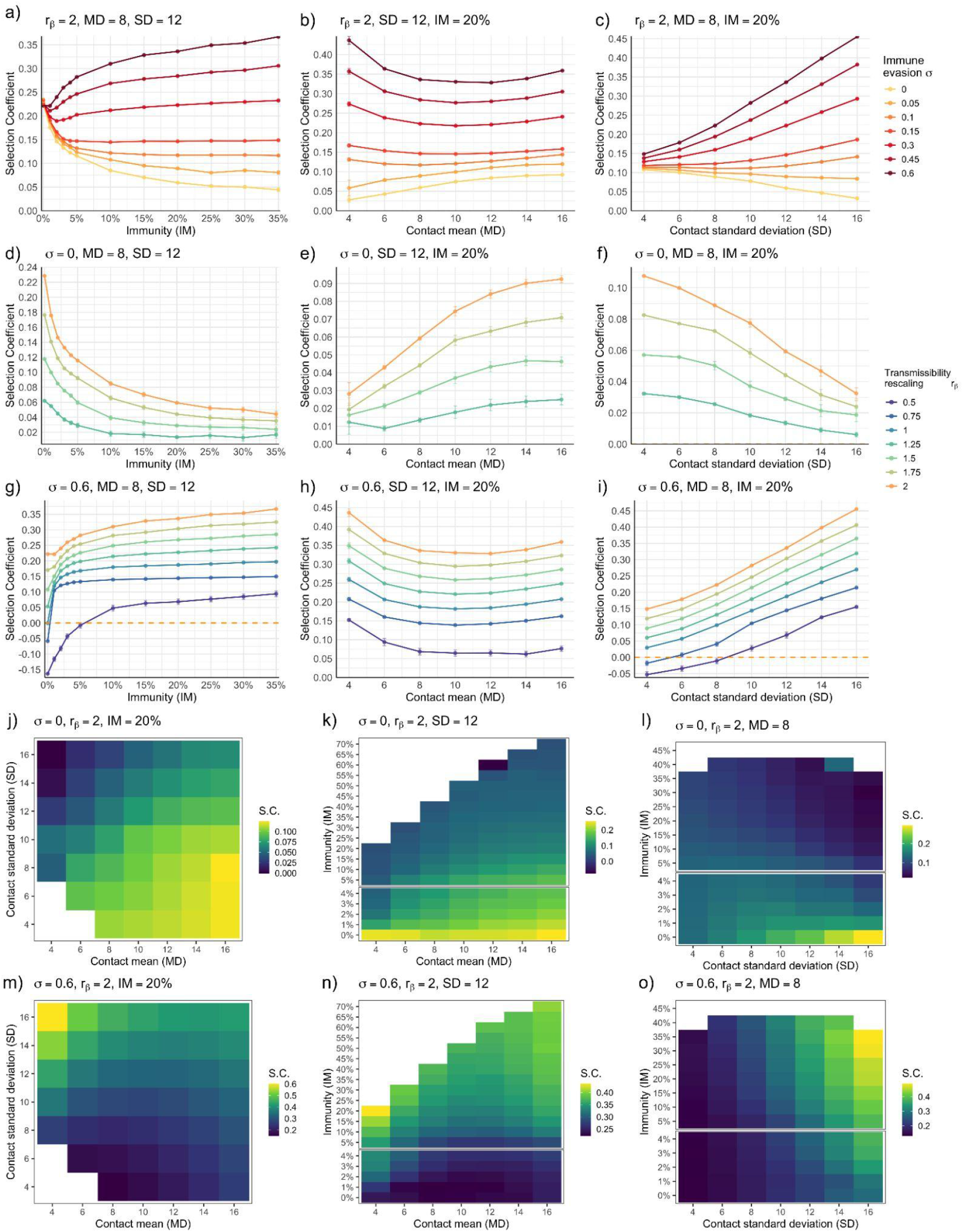
Impact of population characteristics on the selection coefficient of the emerging variant. Panels a – c show the selection coefficient as a function of the immunity (IM) at the time of emergence, mean (MD) and standard-deviation (SD) of contacts, respectively, with the transmissibility rescaling of the emerging variant fixed at *r*_β_ = 2. Colours indicate immune evasion (σ). Fixed parameter values are: (a) MD = 8 and SD = 12; (b) SD = 12 and IM = 20%; (c) MD = 8 and IM = 20%. Panels d – i show the selection coefficient as a function of IM, MD, and SD, respectively, for a fixed immune evasion, where colours represent different transmissibility rescaling factors of the emerging variant (*r*_β_). Panels d-f correspond to σ = 0 (no immune evasion), while g – i to σ = 0.6. Population characteristic parameters, when kept fixed, take the same values as panels a – c. Dots are the mean and the error bars are the standard errors (smaller than the size of the dots for almost all cases). Panels j and m show the heatmap of the selection coefficient as a function of MD and SD, with IM = 0.2. Panels k and n explore MD and IM at fixed SD=12, while panels l and o explore SD and IM at fixed MD=8. Panels j – l correspond to σ = 0, while m – o to σ = 0.6. In the heatmaps, some triplet IM, MD, SD are left white as they could not be explored. The emerging variant could not be introduced at that IM value as this is above the final attack rate of the resident variant for the corresponding MD and SD. The breaks in panels k, l, n, and o separate the low IM values (IM < 5%), sampled at 1% intervals, from the higher IM values. Other parameters are: transmissibility of the resident *γ* = 0. 143 (1/7) *day*s variant β = 0. 0175, recovery rate _−1_, and size of the network *N* = 10^4^. Statistics were computed over 100 stochastic epidemic realisations for each of the 40 stochastic generations of the network with fixed parameters, for a total of 4000 runs. For panels d-f statistics 400 realisation instead of 100 were runned for each network to reduce noise.

Fig. 3b shows that, for σ < 0. 1, the selection coefficient monotonically increased with the contact mean, MD. This was also true for a variant with higher immune evasion, but only for high MD. The non-monotonic trend for σ ≥ 0. 1 mirrors the change of the basic reproductive ratio, which is non-monotonic with MD (see Methods). Consistently, Fig S3 shows that the selection coefficient is associated with the reproductive ratio unless σ is near-zero. This is in agreement with previous works associating the selection coefficient with the reproductive ratio and also with works associating the selection coefficient with the contact mean, as these works implicitly assumed Poisson contact distribution [22].

The selection coefficient, as a function of SD, for fixed MD and IM, is illustrated in Fig. 3c. An increase in contact dispersion slowed down a variant with zero or near-zero immune evasion, while accelerating an immune-evading one. Contact dispersion was found to have a detrimental role on new variant emergence in previous studies that were focusing on the probability of emergence [8,15,17]. Here, we found a similar effect on the selection coefficient. This behaviour can be attributed to the effect of hubs. Hubs can act as super-spreaders or super-blockers. Given their high number of connections, hubs are among the first to be infected, further transmit the infection to a large number of individuals, and are then among the first to become immune. When a variant with no immune evasion emerges, hubs that are already immunised will act as super-blockers. Conversely, a variant able to reinfect immunised hubs can take advantage of their super-spreading role. Of note, we found that the super-blocker effect was fragile and occurred only for no or very low immune evasion, i.e. below σ = 0. 1 for transmission rescaling *r*_β_ = 2 (Fig. 2 c) or even lower when *r*_β_ was smaller (Fig. S4). For σ = 0.05, for instance, the decline with SD occurred only for *r*_β_ > 1. 5 (Fig. S4).

Fig. 3d–i displays how the trends of the selection coefficient in varying IM, MD, and SD change with *r*_β_. In Fig. 3d–f we considered σ = 0, while in Fig. 3g–i σ = 0.6. Overall, the selection coefficient increased with *r*_β_. However, the trends described in Fig. 3a-c remained the same. For the case σ = 0.6 we also explored *r*_β_ < 1 corresponding to the case in which the immune evasion advantage was compensated by a reduction in transmissibility. In this case, we observed that the selection coefficient crossed zero, taking either negative or positive values. The same variant traits either favoured or disfavoured spreading according to population characteristics. In particular, when population immunity was high enough, even a variant with a significant reduction in transmissibility exhibited a greater growth compared with the resident variant. For instance, for *r*_β_ = 0. 5 the selection coefficient became positive when IM was greater than 5% (Fig. 3g).

We conducted a systematic exploration of the parameters IM, MD, and SD, comparing the case of no immune evasion (σ = 0) (Fig. 3j–l). with high immune evasion (σ = 0.6) (Fig. 3m–o). For σ = 0, the increase of the selection coefficient with MD was consistent across all values of immunity and SD tested (Fig. 3j). The decline of the selection coefficient vs. SD, instead, happened only when immunity was sufficiently high. For low values of immunity (below 10%, with all other parameters as in the figure), the trends became non-monotonic (Fig. 3l). For σ = 0. 6 (Fig. 3m–o), the non-monotonic trend of the selection coefficient vs. MD was observed across all IM and SD tested, with the values of MD in correspondence to the minimum selection coefficient changing in varying SD - consistent with the association between the selection coefficient and the reproductive ratio of Fig S3. Despite the selection coefficient being the main focus of our study, we analysed in Fig. S5 the absolute growth of the emerging variant for completeness. We found, in most cases, supercritical spread (see Fig. S5). However, for certain parameter values, the spread remained subcritical, with a negative growth rate. This is consistent with the fact that a variant can still be advantaged over the resident one (i.e., have a positive selection coefficient) even when its spread is subcritical.

We conducted a series of sensitivity analyses to verify the robustness of our results. First, we tested whether the choice of seeding both variants by infecting 50 random individuals could affect the selection coefficient. In Fig. S6, we compared the baseline results of Fig. 3a–c with those obtained using 5 and 10 seeders, showing that the selection coefficient varied slightly for low IM (Fig. S6a), and that such small differences did not affect the overall trends. Still, the number of seeders affected stochastic fluctuations as shown by the fact that the probability of having a positive selection coefficient diminished when the number of seeders was smaller.

We also tested whether other factors potentially present in real situations could alter our results. In Fig. S7, we show that when the emerging variant had, in addition to changes in transmissibility and/or immune evasion, a different generation time, the trends of Fig. 3a–c remained unchanged. In Fig. S8, we considered the effect of spatial coupling. In the baseline scenario, we assumed a well-mixed population and ignored the continuous introduction of both the emerging and the resident variants through travel which could affect the selection coefficient and alter the trends of Fig 3 [48]. To relax this assumption, we simulated the spread of both variants on two contact networks coupled with mobility and modelled the continuous introduction of cases from a second network into the focal one. The characteristics of the second population, IM_2_, MD_2_, and SD_2_, were allowed to differ from those of the focal network – see Supplementary Information for more details. Fig. S8 shows that the trends of the selection coefficient as a function of IM were robust across all tested scenarios. The trends as a function of MD and SD, however, deviated from the baseline in a few cases, particularly when σ was 0, the traveling probability 10^-2^ or above, and MD_2_ was higher than MD.

### Analysis of Alpha emergence across US states

We now consider the Alpha spread in the US as a case study and analyse the relationship between the selection coefficient, contact structure and population immunity. Data on seroprevalence [49], vaccination [50], and contact mean and standard deviation [51] were gathered for all US states during the period of SARS-CoV-2 Alpha emergence and growth, between late December 2020 and late April 2021 (see Methods).

Fig. 4a and b show the evolution in time of the contact mean and standard deviation [51]. They both increased during the study period and varied substantially between states. We defined the set of covariates MD and SD as the averages over time of the mean and standard deviation, respectively, for each state. Their values ranged between 4.1 and 10.9 (mean value 8.0) for MD and between 8.5 and 15.2 (mean value 13.0) for SD. By combining seroprevalence, vaccination data, and vaccine effectiveness we estimated population immunity at the time of Alpha’s emergence for each state, which defined the IM covariate. The mean value across states was 18.6%. Immunity estimated for Vermont was extraordinarily low (1.1%), as the state is rural, with limited connections with urban hubs, and had a strong public health response. On the other hand, the highest immunity was estimated for Ohio and was as high as 30.7%. In Fig. 4 c and d, we show the distribution of MD, SD and IM, as well as their correlation. MD was strongly correlated with SD. This may be explained by the fact that the adaptive behavioural response and the public health interventions, which caused spatiotemporal contact variations during COVID-19, affected different settings (e.g. workplace, social events) and thus they likely altered both contact mean and dispersion. MD was also correlated with IM, likely due to the relatively stable ranking of states according to contact structure: states with higher mean contact likely experienced more transmission, leading to more cumulative infections and, thus, greater built-in immunity by the time Alpha was introduced.

**Fig 4.**
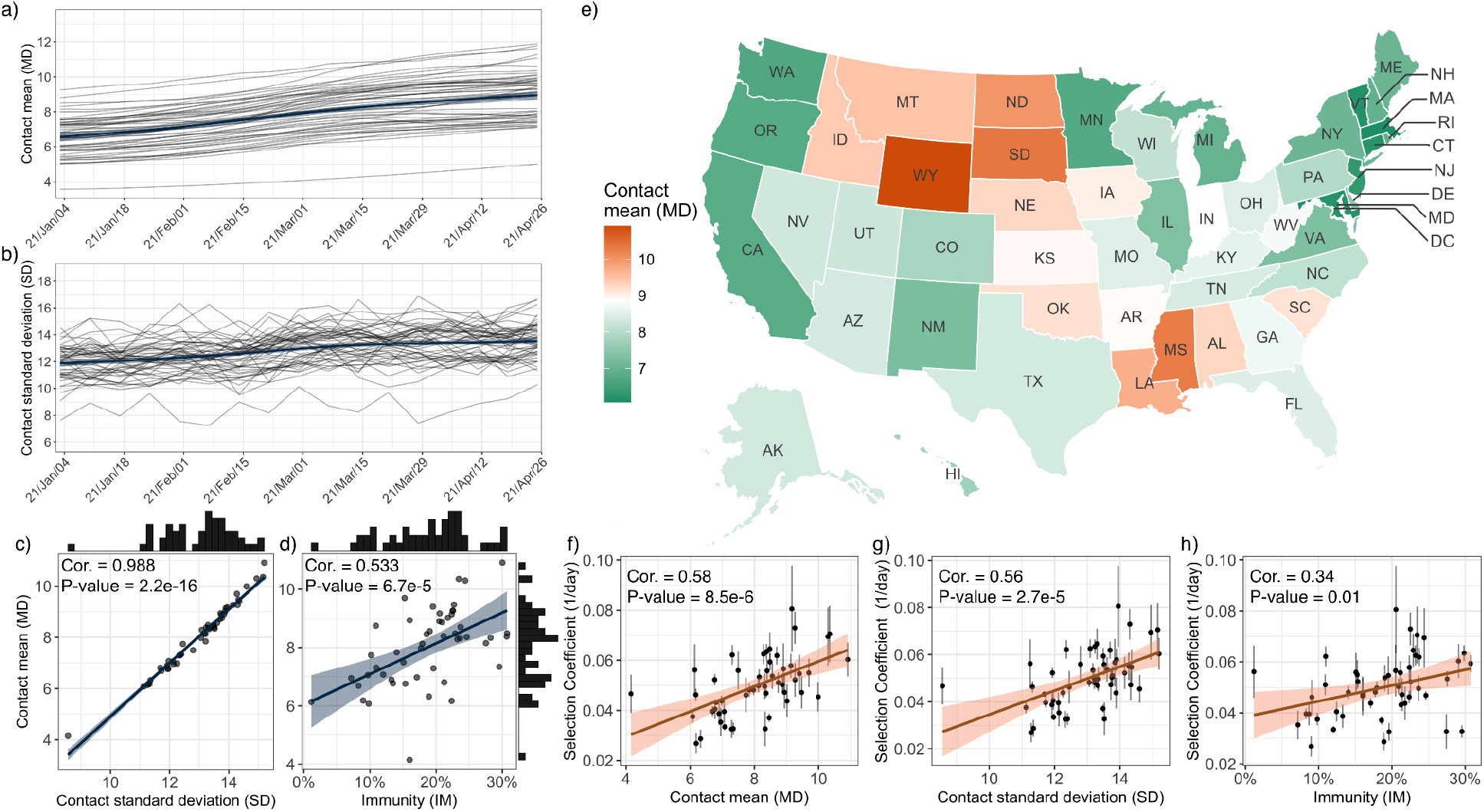
Data from the Alpha variant emergence in the US and statistical analysis. a) Time series of the contact mean, and b) time series of the contact standard deviation. Black curves represent the value of each state over time, and the blue curve is the smoothed curve of the average value. c) Scatter plot of the time-averaged contact mean (MD) against the time-averaged standard deviation (SD) during the period of Alpha emergence, together with their histograms. d) Scatter plot of the time-averaged contact mean (MD) during the period of Alpha emergence and the estimated population immunity (IM) at the time of the Alpha variant’s emergence, together with their histograms. e) Color map of MD for each US state, where white corresponds to the national mean. Base maps are generated using the R package usmap [45] based on geographic boundary data from the U.S. Census Bureau. Figures f-h show the scatter plots of the fitted selection coefficients depending on the population characteristics of each state: f) selection coefficient and MD, g) selection coefficient and SD, h) selection coefficient and IM. The selection coefficient was fitted over the same period used for defining MD and SD.

The map of Fig. 4e allows for a visual comparison of the contact mean by state with the state’s selection coefficient of Fig. 1 b, highlighting that the two quantities have a similar geographical pattern. For instance, all states on the West Coast and many states on the East Coast (especially the Northeast) had both selection coefficient and contact mean below the average. On the other hand, Oklahoma, Nebraska, South Dakota, Arkansas, Louisiana, and Mississippi were among the states with both selection coefficient and contact mean above the average.

Beyond the visual comparison, the scatter plots in Figure 4f–h show that the selection coefficient was positively correlated with all three covariates. Its correlation with MD (Pearson correlation 0.58, *p* = 8. 5 × 10^−6^) and SD (Pearson correlation 0.56, *p* = 2. 7 × 10^−5^) was substantially higher than the one with IM (Pearson correlation 0.35, *p* = 0. 01). Computing the selection coefficient and covariates over the entire window of Alpha emergence could average out temporal variations in the spreading context and hide meaningful interplay between such temporal variations and variant traits [52,53]. We thus repeated the analysis using a 4-week sliding window spanning the entire emergence period, recomputing the selection coefficient, covariates, and correlation values for each window. The positive correlations with MD and SD were overall robust, while that with IM was less stable (Fig. S9, Supplementary Information). We also investigated whether recovered trends could be biased by continuous case introductions from other states. We computed cross-state commuting probabilities from [54] and all-purpose travelling probabilities from the Safegraph Social Distancing dataset [55]. State pairs with high mobility coupling with other states often exhibited similar contact structure (e.g. states in the Northeast). The District of Columbia exhibited a disproportionately high cross-state mobility, likely owing to its size. Excluding it from the correlation analysis slightly increased the correlations with MD and SD (see Supplementary Information). Excluding additional highly connected states did not affect substantially the results for MD and SD, while results for IM were less robust. Overall this suggests that contact structure and immunity could affect the selection coefficient. We used ridge multivariate regression to disentangle the relative role of the three covariates while accounting for collinearity (see Methods). The coefficients obtained identified a stronger effect of MD (5.26×10^-2^), followed by SD (4.96×10^-2^), and IM (2.87×10^-2^). This trend was consistent across penalization approaches and when comparing the explained variance in univariate models.

### Consistency between empirical evidence and theoretical findings

The transmissibility increase of Alpha is estimated at around 50% (*r*_β_ = 1. 5) [42,43,56]. Virological studies reported some, though limited, immune evasion for Alpha [37]. A small reduction in vaccine efficacy was observed [57]. Protection against Alpha from prior infection by the historical variant was estimated at 90% [58]. Another study, however, found no evidence that reinfections were more frequent with Alpha than with the pre-existing variant [59]. These findings are compatible with no or very low immune evasion in our model.

Within this parameter range, the theoretical model predicts an overall positive association between the selection coefficient and MD, which is consistent with the empirical trend. Model predictions regarding the selection coefficient’s association with SD could also be compatible with observations if we assume low but non-zero immune evasion for which the negative effect of SD is absent. To test the consistency between the model and the data, in this parameter regime, we ran simulations with our model with as input the triplets of MD, SD, and IM values estimated for each US state, along with the average household size of each state. We then took transmissibility rescaling *r*_β_ = 1. 5, and immune evasion σ = 0. 1. The simulated and empirical values of the selection coefficient across states were positively correlated (Pearson correlation 0.55, *p* < 10^−4^, after removing Vermont, which was an outlier, Figure S10). In line with the data, the simulated selection coefficient showed positive trends with all covariates, MD, SD, and IM – Pearson correlations 0.84, _−4_, 0.81, *p* = 10^−4^ *p* < 10, and 0.4, *p* < 0. 02, respectively. All correlation values slightly improved when further removing the states with a high volume of introductions from other states as discussed in the previous section (see Supplementary Information). Simulations with lower values of immune evasion, i.e. σ = 0.05, were also correlated with empirical trends. When we set to σ = 0, instead, the model failed to reproduce the empirical trends, and its outputs were not correlated with the data.

## Discussion

The COVID-19 pandemic has brought to the forefront the consequences of viral evolution for epidemic preparedness. The emergence of new variants with distinct epidemiological traits has created a rapidly shifting landscape, challenging both anticipation and the planning of interventions. Understanding the drivers of variant emergence dynamics is thus crucial, not only for anticipating long-term viral evolution, but also for the short-term outbreak response. Theoretical investigations have long explored the relationship between host population characteristics, variant emergence, and pathogen evolution. Classical, decades-old population genetics studies established that the effect of spatial structure on allele fixation is complex [23–26]: spatial mixing can influence the success of an emerging variant, and spatial fragmentation often hinders beneficial mutations, but can also favour them depending on the circumstances [24–26]. This composite view also applies to host-pathogen systems. Prior research has demonstrated that the same host features can have opposite consequences on variant emergence. However, previous findings were obtained with diverse model settings and different metrics for variant advantage, preventing a clear understanding of how the system transitions from one regime to another. The consistent exploration of different ingredients within the same model, which explicitly accounts for the mechanisms underlying the infection dynamics, allows for interpretable results that can be put in relation to empirical data. Our contribution to this research effort was twofold. First, we carried out such an exploration with a model tailored for acute respiratory infections on a well-mixed population and carried out extensive numerical reconstruction of the possible dynamical trends in varying host characteristics and pathogen traits. Second, we analysed real-world data to show that empirical patterns are consistent with theoretical expectations.

We focused on two variant traits – change in transmissibility and immune evasion – and three host population characteristics – immunity, contact mean and dispersion. We designed a spreading model to capture the effect of these ingredients, maintaining a parsimonious and general description of the disease dynamics to provide an understanding of the basic ecological behaviour. This allowed us to better characterise the boundaries of previously identified dynamical regimes. We found that increased contact dispersion hindered variant emergence only in a narrow parameter region, namely, near-zero immune evasion and only when both immunity and the increase in transmission were sufficiently large. Otherwise, the selection coefficient increased with the reproductive ratio, which for heterogeneous uncorrelated networks is a non-monotonic function of contact mean, when contact mean was low, becoming monotonic at higher contact mean. The effect of immunity diminished as immunity increased. For variants with immune evasion and reduction in transmissibility, changes in population immunity determined a shift between a regime of negative selection coefficient and positive one.

Univariate analyses of Alpha emergence across U.S. states showed that the selection coefficient correlated with contact mean and dispersion as well as with population immunity, suggesting that host population characteristics could play a role in variant dynamics. To account for strong correlations among predictors, particularly between the mean and standard deviation of contacts, we applied multivariate ridge regression, which confirmed positive associations with all three factors, though weaker for immunity. Correlation with immunity was also less robust in our sensitivity tests. Model simulations, parameterized with Alpha-specific traits and state-level characteristics, reproduced these empirical patterns and showed agreement with observed trends, under the hypothesis of limited albeit non-zero immune evasion, which could be compatible with virological findings.

Overall, results suggest an important role of contact structure. A previous study reported a negative association between the stringency of interventions and the selection coefficient across regions in the UK [33]. Considering that interventions reduce contact rates, our results are therefore consistent with that work. Here, however, we directly investigated the role of contacts, which, while influenced by interventions, are not solely determined by them, as factors such as adherence and adaptive behavioural response also play a role [51,60–62]. The effect of immunity appeared weaker than that of contact patterns. Differently from Alpha, Omicron was marked by strong immune evasion, and a stronger impact of immunity would therefore be expected. Future work could extend the present analysis to Omicron emergence, leveraging model-based extrapolations of human contact structure that have been validated in previous studies [51]. However, the complexity of the immunity landscape and the greater change in epidemiology compared to Alpha, would complicate this analysis.

Human face-to-face contacts are important drivers of respiratory infection dynamics. These are, however, difficult to reconstruct from data. Over the last two decades, extensive efforts have produced either detailed, high-resolution records of contact dynamics in specific settings, for small populations, and over short time periods [63–67], or coarse contact statistics of daily interactions obtained from large representative samples of the population at national scales [51,68–70]. These data sources provide a fragmented picture of human contact patterns, which complicates epidemic assessment—particularly in situations such as the COVID-19 pandemic, where adaptive behavioural responses and public health interventions caused abrupt changes in these patterns. The COVID-19 pandemic has intensified efforts to reconstruct contact structure and, more broadly, human behaviour, and has opened a reflection on essential information that needs to be collected for modelling [51,70,71]. Here, we show that, in addition to contact mean, their dispersion also plays an important role in determining the dynamics of variant emergence, yet this indicator is rarely investigated in survey studies. It is important to note that to integrate this information into models, a challenge arises from the nature of surveys, which typically record the number of contacts on a single day and thus cannot capture the temporal dynamics of contacts on timescales relevant for infection spread.

While we focused on traits subject to selection in emerging respiratory infections — ranging from influenza to SARS-CoV-2 — other traits were left unexplored, as including them would render the parameter space exceedingly vast. First, we did not explore differences in generation time between variants. Previous studies have shown a complex interplay between generation time and the timescale of host mixing (where mixing refers to both contact patterns and mobility, depending on the scale of analysis): high mixing favours pathogens with shorter generation times, while low mixing favours those with longer ones [16,52,72,73]. While we did not address these dynamical effects here, we performed a sensitivity analysis to ensure that changes in generation time (infectious period in our model) did not affect our conclusions. For SARS-CoV-2 specifically, given the substantial changes in contact means driven by social restrictions, it has been suggested that such dynamical interplay with generation time could play a role for variants with altered infectivity profiles [52]. This is unlikely to have affected our Alpha case study, since the majority of studies agree that the generation time of Alpha was consistent with that of the historical strain [52,53,74]. Moreover, our sliding window analysis showed that correlation results remained broadly stable over time, suggesting that no complex dynamical effects linked to shifts in the epidemiological context affected our findings.

Second, we did not explore changes in virulence. Variations in disease severity may alter transmission and affect the selection coefficient in multiple ways. Higher symptom severity is often associated with greater transmissibility, suggesting that more severe variants could also be more transmissible. At the same time, more severe infections may be more readily detected through increased hospitalisation and reduced asymptomatic spread, potentially facilitating control and effectively shortening the infectious period. Virulence has therefore been extensively studied in relation to spreading timescales, yielding results broadly analogous to those observed for generation time [14,22,33,73,75].

Our study focuses on variant frequency and the selection coefficient as the primary metrics for measuring variant advantage, assuming constant logistic growth throughout the emergence period. More sophisticated approaches have recently been developed for real-time variant growth evaluation and risk assessment, capable of handling scarce genetic data and reporting delays; by focusing on absolute variant growth and reproductive ratio, these offer better risk assessment capabilities [7,76,77]. Still, while the constant logistic growth assumption fits the data only imperfectly, it provides a simple yet meaningful indication of average relative epidemic speed that can be linked to population characteristics, effectively bridging the gap between theoretical and data-driven analyses. The stylised model developed here was not designed to capture the full complexity of the COVID-19 epidemic across US states or to achieve agreement with absolute incidence growth. One aspect that challenges the quantitative match between theoretical and empirical analyses is the potential bias introduced by using survey-based contact structure as input to a static network model. Variations in contact numbers observed across individuals on a given day may reflect short-term fluctuations rather than stable behavioural differences. Daily survey data may therefore overestimate heterogeneities that would average out over longer timescales. This was previously found to bias projections on the herd immunity threshold [78–82]. The same effect would limit here the ability of the model to simultaneously capture the behavior of the selection coefficient and incidence, which is beyond the scope of the work. The fact that the simulated selection coefficient nonetheless correlates with empirical observations, under reasonable assumptions, indicates that the qualitative behaviour described here is robust to many dynamic complexities not explicitly represented in the model.

We considered a locally well-mixed population, albeit with heterogeneous contact structure. Epidemics are multiscale phenomena [83], and regional source-sink dynamics proved central to the early seeding of new variants [43,84–86]. For Alpha in the UK, a unidirectional flow of introductions from Greater London to other regions was initially observed, which may have biased early estimates of Alpha’s selection coefficient in those regions [84]. As local outbreaks grew, however, importations from the source became progressively less important [84]. Alpha’s introduction in the US followed a multicentre pattern, with several states fuelling regional spread between November and January [41,42]. Our study focuses on a later stage, from late January to late April, when Alpha was already spreading locally across all states included in the analysis. In this regime, local cases typically dominate importations unless the traveller flux is particularly strong [48]. Our sensitivity analysis showed that local selection coefficient trends in the simulations were perturbed when the incoming traveller fraction exceeded ~1% and originated from a more connected population, i.e. with greater transmission potential. With few exceptions, cross-state mobility fractions remained below this threshold, and stronger fluxes typically connected states with similar contact patterns (e.g. Northeast states). A notable exception was Washington, DC which exhibits disproportionately high cross-state mobility. Removing Washington, DC from the correlation analysis yielded a slight improvement.

This study is subject to limitations. First, we modelled immunity in a polarised way, neglecting heterogeneous immune evasion and homologous reinfection. Accounting for heterogeneous immune evasion requires complex analyses that go beyond the scope of this work. Neglecting homologous reinfection, on the other hand, is acceptable over short time scales, when infections due to the resident variant are recent. Indeed, homologous reinfection was found to be rare during the first months of COVID-19 spread, including the period of Alpha emergence [59]. Similarly, homologous reinfection of influenza within the same season has been shown to play a limited role in transmission [87]. Second, we did not account for behavioral factors other than contacts, such as use of masks and adoption of hygiene measures. Also we could not distinguish between indoor and outdoor contacts. Third, while we investigated contact heterogeneities, the model does not incorporate the socioeconomic and demographic drivers of such heterogeneities—e.g., age, education, income—which have been shown to shape patterns of COVID-19 spread [51,88–90]. Explicitly accounting for their role in variant dynamics is an important future direction. Finally, our analysis of Alpha spread across the US was conducted at the state level. Differences in contact structure across states were largely driven by marked differences in urbanisation [51]. Analyses at higher spatial resolution could better account for this factor and reduce noise; however, sequencing coverage was insufficient to support analyses at finer scales.

This study demonstrates that contact networks and immunity are associated with the selection coefficient of emerging pathogen variants, with contact dispersion playing opposing roles depending on a variant’s immune evasion capacity. From a public health perspective, these findings suggest that surveillance and intervention strategies should be tailored to local population characteristics, with contact reduction measures potentially having differential effects on variant competition. Understanding these dynamics could inform targeted vaccination strategies and enhance variant emergence prediction when integrated with real-time behavioral surveillance systems that capture evolving contact patterns.

## Methods

### Selection coefficient

The ecological dynamics of the emerging variant can be reconstructed by looking at its frequency, *p*(*t*), i.e. the percentage of incidence cases attributed to it: 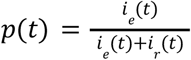, where *i*_e_(*t*) and *i*_r_(*t*) are the incidences of the emerging and the resident variant, respectively. The selection coefficient, *s*(*t*), measures the growth of the emerging variant frequency relative to the resident variant one,

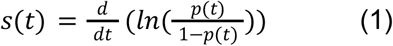

When *s*(*t*) is constant in time, the emerging variant frequency *p*(*t*) follows a logistic function,

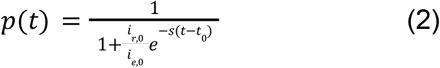

where *i*_*r*,0_ and *i*_*e*,0_ are the incidences of the emerging and the resident variant, respectively, evaluated at time *t*_0_. According to equation (2), the frequency *p*(*t*) increases if *s* > 0 and decreases when *s* < 0. We estimate the selection coefficient by fitting the logistic function *p*(*t*) to the frequency curve, for both the simulated and real frequency - see below for details on the definition of the time window for the fit. We regress a binomial Generalized Linear Model (GLM) with a logit link function in the R programming language to estimate the parameters of the logistic function and their confidence intervals.

### Computational model of two-variant infection dynamics

#### Modelling human-to-human contact network

We model a two-layer contact network with N individuals (nodes). The first layer accounts for household contacts, while the second layer accounts for non-household contacts (e.g. occurring in the workplace, school or community). To construct the household layer, we distribute the nodes in households of different sizes and consider a clique, i.e. a fully connected graph, inside each household. We assume that the household size follows a zero-truncated Poisson distribution [91], 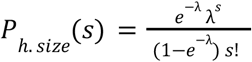. This leads to a Poisson distribution for the nodes’ degree in the household layer, 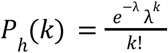, where the subscript *h* indicates the household layer.

For the non-houseshold layer, that we label *n. h*., we assume that the probability of an individual having a degree *k* follows a negative binomial distribution shifted by 1 (to avoid agents with no contacts in this layer), 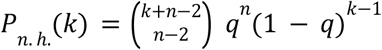, with *k* ≥ 1 and free parameters *n* and *q*, that are related to the mean degree and the standard deviation of the degree as follow: 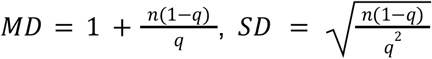. Once assigned a degree to each node based on this distribution, we connect the stubs using the configuration model to avoid degree-degree correlations.

We parametrised the networks as follows. The network size was *N* = 10^4^, a compromise between being reasonably large and maintaining computational times to feasible levels. We set the household-size distribution parameter to λ = 2, yielding an average household size of 2.3 – similar to values observed in European countries and the US [92,93]. We then varied the parameters of the non-household layer to explore MD and SD within ranges estimated from US (see Fig. 4) and compatible with European surveys as well – namely, MD in [4, 16] and SD in [3, 16].

### Modelling transmission

We use a susceptible-infected-recovered (SIR) model to simulate the spread of two variants in the network. In this model, the population is distributed into the compartments, S (Susceptible), I (Infected), and R (Recovered), with respect to each variant. We assume that co-infection is not possible, i.e. no individuals can be infected with more than one strain at the same time. Therefore, Individuals are divided into eight compartments: fully susceptible (*S*), infected by the resident variant (*I*_r_), infected by the emerging variant (*I_e_*), recovered from the resident variant and still susceptible to the emerging variant (*R*^(*r*)^), recovered from the resident variant and infected by the emerging variant (*I*^(*r*)^*_e_*), recovered from the emerging variant and still susceptible to the resident variant (*R*^(*e*)^), recovered from the emerging variant and infected by the resident variant (*I*^(*e*)^ _*r*_), and recovered from all variants (*R*). Susceptible agents can be infected by their infectious neighbors with transmission rates per contact β_*r*_ and β_*e*_ for resident and emerging variants, respectively. We quantify the change in transmission of the emerging variant with the rescaling parameter 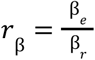. The cross-immunity parameter *σ_j,i_* represents the reduction in susceptibility to the variant *i* due to the acquired immunity to *j*. Namely, individuals previously infected by the variant *j* can contract the infection by *i* with the per-contact rate σ_j,i_ β_i_, where *σ_i,j_* = 0 and *σ_i,j_* = 1 represent full cross-immunity and no cross-immunity, respectively. We assume the symmetry condition applies, i.e. *σ*_*i*,*j*_ = *σ*_*j*,i_ = *σ*, as depicted in Fig. 2 b. Infectious individuals recover with a rate *γ*, assumed to be the same for the two variants.

In the supplementary information we present two sensitivity analyses where *a)* we considered the case in which recovery rates of the emerging variant is different from the resident one (in Fig. S7), and *b)* assume a metapopulation where the individuals are divided in two networks coupled through mobility, to address the role of continuous introductions from the outside in shaping selection coefficient trends (section “Impact of mobility between populations” of the supplementary information and Fig. S8).

The transmission model was parametrised to consider a scenario similar to the emergence of a SARS-CoV-2 variant. The recovery rate was set to *γ* = 1/(7 *day*s). The per-contact transmission rate of the resident variant β_r_ was set to 0. 0175 per day. The basic reproductive ratio of the resident variant corresponding to the input MD and SD can be computed from the equation 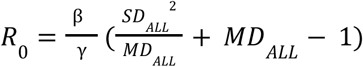, where MD_ALL_ and SD_ALL_ are contact mean and standard deviation of the whole network composed by both the household and non-household layer. The explored SD and MD values lead to *R*_0_ values within the range [0.98 - 4.82]. Variant’s change in transmission and immune evasion were varied, considering values of *r*_β_ between 0.5 and 2 and σ between 0 and 0.6.

### Details of the numerical simulations

Simulations are stochastic and discrete-time. At each simulation time-step, corresponding to one day, we simulate the two processes:

-Infection: Every agent infected with the variant *i* can transmit the infection to neighbouring nodes that are susceptible to *i* with probability β_*i*_ or σ_*i*,*j*_ β_*i*_, according to whether they have no infection history or a history of *j* infection, respectively.

-Recovery: Every infectious individual recovers from the infection with a probability *γ*.

To simulate the spreading of the variants, we initially infect 50 random individuals with the resident variant and set the state of the rest of the population to *S*. The value of 50 was chosen to avoid the initial epidemic extinction from occurring too often. Then we let the simulation continue until the time when the proportion of recovered by the resident variant reached IM. At this point, the emerging variant is seeded by randomly infecting 50 susceptible individuals. The simulation continues until no infectious agents remain.

For each set of parameters of the contact network, we generate an ensemble of 40 random networks. For each generated network, we then run 100 stochastic realizations of the epidemic. For certain parameter values it was necessary to run more simulations (400 instead of 100) to reduce noise.

For each stochastic run, we computed the selection coefficient by fitting the logistic curve to the emerging variant frequency (see the dedicated section at the beginning of the Methods section). The time window used for the fit was defined from the emergence of the variant until either the time of the incidence peak or 7 days after the emergence, whichever occurs later. In the Supplementary Information we also analyzed the growth rate of the emerging variant’s incidence. The growth rate at time step *t* is defined as *G*(*t*) = ln(*i_e_*(*t* + 1)) −ln(*i_e_*(*t*)), where *i_e_*(*t*) is the incidence of the emerging variant. We computed the time-averaged growth rate as the average *G*(*t*) over seven days from the emergence day.

Both the growth rate and the selection coefficient were computed for each stochastic run. Figure 2, 3 and other figures in the SI show statistics over all runs.

### Empirical analysis of Alpha spread across US states

#### SARS-CoV-2 Alpha’s frequency and selection coefficient

The data used to estimate Alpha frequency were provided by the CoV-Spectrum project [44]. The Alpha variant took about 90 days to reach its peak incidence in each state. Therefore, we defined the time window of Alpha growth as the period with a length of 12 weeks (84 days) centred around the midpoint date when the Alpha frequency reaches half of its maximum value. In the cases in which Alpha was peaking and starting to decline before the midpoint plus six weeks, the date of the peak frequency was taken as the end of the window (Figure S1).

#### Face-to-Face contact structure

For all US states, the number of non-household contacts was obtained from the U.S. COVID-19 Trends and Impact Survey (Delphi Research Group at Carnegie Mellon University in partnership with Facebook). The dataset is described and analysed in [51]. Briefly, in the survey, contact was defined as a face-to-face conversation or physical contact. Contacts are aggregated on a weekly basis for each U.S. state, and the mean and standard deviation over the number of contacts are computed after removing outlier responses with the number of contacts above the 95th percentile. The data spans the period from April 27, 2020, to April 26, 2021.

Weekly mean and standard deviation in the number of contacts are then averaged during the time window of Alpha growth defined above to obtain the covariates, MD and SD.

#### Population’s immunity

For each US state, we estimated the effective population immunity against SARS-CoV-2 resident variant using seroprevalence [49] and vaccination [50] data provided by the Centers for Disease Control and Prevention (CDC). Assuming *z*(*t*) is the proportion of the population infected before the time of Alpha emergence, *v*^*p*^ (*t*) is partial vaccination coverage (i.e. one dose of a two-dose vaccine schedule) and *v*^*c*^ (*t*) is complete vaccination coverage, then effective immunity can be estimated as [94]:

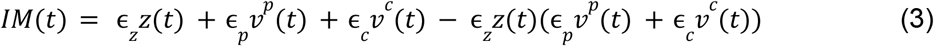

Where ϵ_*z*_, ϵ_*p*_ and ϵ_*c*_ are natural immunity efficacy, partial vaccination efficacy, and complete vaccination efficacy, respectively. We used the following values informed by the literature [94]: ϵ_*z*_ = 1, ϵ_*p*_ = 0. 6 and ϵ_*c*_ = 0. 9.

The covariate IM of each state used in the regression analysis below was defined as *IM*(*t*), where *t* is the starting date of the window of Alpha growth. For North Dakota, data were partial and it was not possible to compute the immunity, therefore it was not included in the correlation analysis where IM was involved.

#### Regression analysis

To disentangle the association of MD, SD, and IM with the selection coefficient (S), we performed ridge regression. Ordinary least squares regression can be unstable in the presence of multicollinearity (as observed between MD, SD, and IM) and ridge regression addresses this issue by imposing an L_2_-norm penalty on all model coefficients, shrinking them towards zero while keeping them in the model. We used a Gaussian family and z-normalized all covariates. We performed k-fold cross-validation using the R package glmnet to choose a penalty tuning parameter, λ, which optimizes the bias-variance tradeoff. We select the maximum value of λ so that the mean-squared error is within one standard deviation of the minimum [95]. Our results are qualitatively robust to an alternative choice of λ that minimizes the mean squared error, and other penalty formulations including LASSO L_1_-norm penalties. Standard errors are not reported as confidence intervals do not have usual frequentist properties in ridge regression.

## Data availability

Numbers of SARS-CoV-2 and SARS-CoV-2 Alpha sequences were obtained from the CoV-Spectrum project [44] (https://cov-spectrum.org/) from GISAID data (https://gisaid.org) [96]. These data were publicly accessible before December 2025. Additional information on the data and minimal data to reproduce the study are provided on GitHub at https://github.com/PouryaTS/MultiStrain_Network. Seroprevalence [49] (https://data.cdc.gov/Laboratory-Surveillance/Nationwide-Commercial-Laboratory-Seroprevalence-Su/d2tw-32xv/about_data) and vaccination [50] (https://data.cdc.gov/Vaccinations/COVID-19-Vaccinations-in-the-United-States-Jurisdi/unsk-b7fc/about_data) data were obtained from the Centers for Disease Control and Prevention (CDC). Data on the average household size for each U.S. state were retrieved from Statista (https://www.statista.com/statistics/242265/average-size-of-us-households-by-state/) [97]. The number of non-household contacts was obtained from the U.S. COVID-19 Trends and Impact Survey (Delphi Research Group at Carnegie Mellon University in partnership with Facebook). The number of cross-state commuters between 2016 and 2020 were obtained from the United States Census Bureau (https://www.census.gov/data/tables/2020/demo/metro-micro/commuting-flows-2020.html) [54]. Reduction of mobility in workplaces was obtained from the Community Mobility Reports by Google (https://www.google.com/covid19/mobility/). All-purpose mobility fluxes at the county-day scale in the U.S. was obtained from Safegraph Social Distancing dataset (https://docs.safegraph.com/docs/social-distancing-metrics) [55]. Maps were generated in R using the usmap package [45], which provides U.S. state boundaries derived from the U.S. Census Bureau Cartographic Boundary Files (https://www.census.gov/geographies/mapping-files/time-series/geo/cartographic-boundary.html). The weekly, state-level means and variances of the number of contacts used in this study, number of sequences counts used for the logistic fit, along with scripts for running simulations, generating raw simulation data, and estimating selection coefficients, are publicly available on GitHub at https://github.com/PouryaTS/MultiStrain_Network.

## Supporting information

S1 Appendix. Supplementary text and figures.

## Acknowledgments

We thank Pierre-Yves Boëlle and Mircea Sofonea for useful discussion. We acknowledge financial support by the Municipality of Paris through the programme Émergence(s) to CP and PTS; Cariparo Foundation through the program Starting Package to CP; Department of Molecular Medicine through the program SID from BIRD funding to CP. We gratefully acknowledge support from the ANRS MIE Network on Modelling Infectious Diseases, which provided a travel grant to PTS. for a research visit. The funders had no role in the study design, data collection and analysis, decision to publish, or preparation of the manuscript.

## Author contributions

Conceptualization: EV, LO, SB, CP. Data curation: PTS, JCT, LO, SB, CP. Formal analysis: PTS, JCT, EV, LO, SB, CP. Investigation: PTS, JCT, EV, LO, SB, CP. Methodology: PTS, JCT, EV, LO, SB, CP. Software: PTS. Supervision: SB, CP. Validation: PTS, JCT, LO, SB, CP. Visualization: PTS. Writing – original draft: PTS, CP. Writing – review & editing: PTS, JCT, LO, SB, CP.

## References

1. Matt J. Keeling PR. Modeling Infectious Diseases in Humans and Animals. Princeton University Press; 2008.

2. Pastor-Satorras R, Castellano C, Van Mieghem P, Vespignani A. Epidemic processes in complex networks. Rev Mod Phys. 2015;87: 925–979. doi:10.1103/RevModPhys.87.925

3. Bansal S, Grenfell BT, Meyers LA. When individual behaviour matters: homogeneous and network models in epidemiology. J R Soc Interface. 2007;4: 879–891. doi:10.1098/rsif.2007.1100

4. Valdano E, Colombi D, Poletto C, Colizza V. Epidemic graph diagrams as analytics for epidemic control in the data-rich era. Nat Commun. 2023;14: 8472. doi:10.1038/s41467-023-43856-1

5. Kiss IZ, Miller J, Simon PL. Mathematics of Epidemics on Networks: From Exact to Approximate Models. Springer International Publishing; 2017. doi:10.1007/978-3-319-50806-1

6. Bedford T, Riley S, Barr IG, Broor S, Chadha M, Cox NJ, et al. Global circulation patterns of seasonal influenza viruses vary with antigenic drift. Nature. 2015;523: 217–220. doi:10.1038/nature14460

7. Volz E. Fitness, growth and transmissibility of SARS-CoV-2 genetic variants. Nat Rev Genet. 2023; 1–11. doi:10.1038/s41576-023-00610-z

8. Pinotti F, Fleury É, Guillemot D, Böelle P-Y, Poletto C. Host contact dynamics shapes richness and dominance of pathogen strains. PLOS Comput Biol. 2019;15: e1006530. doi:10.1371/journal.pcbi.1006530

9. Shrestha S, King AA, Rohani P. Statistical Inference for Multi-Pathogen Systems. PLOS Comput Biol. 2011;7: e1002135. doi:10.1371/journal.pcbi.1002135

10. Atkins KE, Lafferty EI, Deeny SR, Davies NG, Robotham JV, Jit M. Use of mathematical modelling to assess the impact of vaccines on antibiotic resistance. Lancet Infect Dis. 2018;18: e204–e213. doi:10.1016/S1473-3099(17)30478-4

11. Wu JT, Leung GM, Lipsitch M, Cooper BS, Riley S. Hedging against Antiviral Resistance during the Next Influenza Pandemic Using Small Stockpiles of an Alternative Chemotherapy. PLOS Med. 2009;6: e1000085. doi:10.1371/journal.pmed.1000085

12. Day T, Kennedy DA, Read AF, Gandon S. Pathogen evolution during vaccination campaigns. PLOS Biol. 2022;20: e3001804. doi:10.1371/journal.pbio.3001804

13. Park SW, Cobey S, Metcalf CJE, Levine JM, Grenfell BT. Predicting pathogen mutual invasibility and co-circulation. Science. 2024;386: 175–179. doi:10.1126/science.adq0072

14. Lion S, Boots M. Are parasites ‘“prudent”‘ in space? Ecol Lett. 2010;13: 1245–1255. doi:10.1111/j.1461-0248.2010.01516.x

15. Leventhal GE, Hill AL, Nowak MA, Bonhoeffer S. Evolution and emergence of infectious diseases in theoretical and real-world networks. Nat Commun. 2015;6: 6101. doi:10.1038/ncomms7101

16. Poletto C, Meloni S, Colizza V, Moreno Y, Vespignani A. Host Mobility Drives Pathogen Competition in Spatially Structured Populations. PLoS Comput Biol. 2013;9: e1003169. doi:10.1371/journal.pcbi.1003169

17. Elie B, Selinger C, Alizon S. The source of individual heterogeneity shapes infectious disease outbreaks. Proc R Soc B Biol Sci. 2022;289: 20220232. doi:10.1098/rspb.2022.0232

18. Reyné B, Djidjou-Demasse R, Sofonea MT, Alizon S. Mutant emergence timing and population immunisation status impact epidemiological dynamics. J Theor Biol. 2025;608: 112140. doi:10.1016/j.jtbi.2025.112140

19. Bushman M, Kahn R, Taylor BP, Lipsitch M, Hanage WP. Population impact of SARS-CoV-2 variants with enhanced transmissibility and/or partial immune escape. Cell. 2021;184: 6229-6242.e18. doi:10.1016/j.cell.2021.11.026

20. Bansal S, Meyers LA. The impact of past epidemics on future disease dynamics. J Theor Biol. 2012;309: 176–184. doi:10.1016/j.jtbi.2012.06.012

21. Fox SJ, Miller JC, Meyers LA. Seasonality in risk of pandemic influenza emergence. PLOS Comput Biol. 2017;13: e1005749. doi:10.1371/journal.pcbi.1005749

22. Day T, Gandon S, Lion S, Otto SP. On the evolutionary epidemiology of SARS-CoV-2. Curr Biol. 2020;30: R849–R857. doi:10.1016/j.cub.2020.06.031

23. Maruyama T. Effective number of alleles in a subdivided population. Theor Popul Biol. 1970;1: 273–306. doi:10.1016/0040-5809(70)90047-X

24. Barton NH. The probability of fixation of a favoured allele in a subdivided population. Genet Res. 1993;62: 149–157. doi:10.1017/S0016672300031748

25. Whitlock MC. Fixation Probability and Time in Subdivided Populations. Genetics. 2003;164: 767–779. doi:10.1093/genetics/164.2.767

26. Cherry JL. Selection, Subdivision and Extinction and Recolonization. Genetics. 2004;166: 1105–1114. doi:10.1093/genetics/166.2.1105

27. Day T, Gandon S. Applying population-genetic models in theoretical evolutionary epidemiology. Ecol Lett. 2007;10: 876–888. doi:10.1111/j.1461-0248.2007.01091.x

28. Berngruber TW, Froissart R, Choisy M, Gandon S. Evolution of Virulence in Emerging Epidemics. PLOS Pathog. 2013;9: e1003209. doi:10.1371/journal.ppat.1003209

29. Lopez S, Komarova NL. An optimal network that promotes the spread of an advantageous variant in an SIR epidemic. J Theor Biol. 2025;605: 112095. doi:10.1016/j.jtbi.2025.112095

30. Lieberman E, Hauert C, Nowak MA. Evolutionary dynamics on graphs. Nature. 2005;433: 312–316. doi:10.1038/nature03204

31. Hindersin L, Traulsen A. Most Undirected Random Graphs Are Amplifiers of Selection for Birth-Death Dynamics, but Suppressors of Selection for Death-Birth Dynamics. PLOS Comput Biol. 2015;11: e1004437. doi:10.1371/journal.pcbi.1004437

32. Yagoobi S, Traulsen A. Fixation probabilities in network structured meta-populations. Sci Rep. 2021;11: 17979. doi:10.1038/s41598-021-97187-6

33. Otto SP, Day T, Arino J, Colijn C, Dushoff J, Li M, et al. The origins and potential future of SARS-CoV-2 variants of concern in the evolving COVID-19 pandemic. Curr Biol. 2021;31: R918–R929. doi:10.1016/j.cub.2021.06.049

34. Boots M, Mealor M. Local Interactions Select for Lower Pathogen Infectivity. Science. 2007;315: 1284–1286. doi:10.1126/science.1137126

35. Benhamou W, Blanquart F, Choisy M, Berngruber TW, Choquet R, Gandon S. Evolution of virulence in emerging epidemics: from theory to experimental evolution and back. Virus Evol. 2024;10: veae069. doi:10.1093/ve/veae069

36. Alizon S, Sofonea MT. SARS-CoV-2 virulence evolution: Avirulence theory, immunity and trade-offs. J Evol Biol. 2021;34: 1867–1877. doi:10.1111/jeb.13896

37. Tao K, Tzou PL, Nouhin J, Gupta RK, de Oliveira T, Kosakovsky Pond SL, et al. The biological and clinical significance of emerging SARS-CoV-2 variants. Nat Rev Genet. 2021;22: 757–773. doi:10.1038/s41576-021-00408-x

38. Brito AF, Semenova E, Dudas G, Hassler GW, Kalinich CC, Kraemer MUG, et al. Global disparities in SARS-CoV-2 genomic surveillance. Nat Commun. 2022;13: 7003. doi:10.1038/s41467-022-33713-y

39. Subissi L, von Gottberg A, Thukral L, Worp N, Oude Munnink BB, Rathore S, et al. An early warning system for emerging SARS-CoV-2 variants. Nat Med. 2022;28: 1110–1115. doi:10.1038/s41591-022-01836-w

40. Campbell F, Archer B, Laurenson-Schafer H, Jinnai Y, Konings F, Batra N, et al. Increased transmissibility and global spread of SARS-CoV-2 variants of concern as at June 2021. Eurosurveillance. 2021;26: 2100509. doi:10.2807/1560-7917.ES.2021.26.24.2100509

41. Alpert T, Brito AF, Lasek-Nesselquist E, Rothman J, Valesano AL, MacKay MJ, et al. Early introductions and transmission of SARS-CoV-2 variant B.1.1.7 in the United States. Cell. 2021;184: 2595-2604.e13. doi:10.1016/j.cell.2021.03.061

42. Washington NL, Gangavarapu K, Zeller M, Bolze A, Cirulli ET, Schiabor Barrett KM, et al. Emergence and rapid transmission of SARS-CoV-2 B.1.1.7 in the United States. Cell. 2021;184: 2587-2594.e7. doi:10.1016/j.cell.2021.03.052

43. Faucher B, Sabbatini CE, Czuppon P, Kraemer MUG, Lemey P, Colizza V, et al. Drivers and impact of the early silent invasion of SARS-CoV-2 Alpha. Nat Commun. 2024;15: 2152. doi:10.1038/s41467-024-46345-1

44. Chen C, Nadeau S, Yared M, Voinov P, Xie N, Roemer C, et al. CoV-Spectrum: analysis of globally shared SARS-CoV-2 data to identify and characterize new variants. Bioinformatics. 2022;38: 1735–1737. doi:10.1093/bioinformatics/btab856

45. Lorenzo PD. usmap: US Maps Including Alaska and Hawaii. 2025. Available: https://cran.r-project.org/web/packages/usmap/index.html

46. Hinch R, Panovska-Griffiths J, Probert WJM, Ferretti L, Wymant C, Di Lauro F, et al. Estimating SARS-CoV-2 variant fitness and the impact of interventions in England using statistical and geo-spatial agent-based models. Philos Transact A Math Phys Eng Sci. 380: 20210304. doi:10.1098/rsta.2021.0304

47. Roquebert B, Trombert-Paolantoni S, Haim-Boukobza S, Lecorche E, Verdurme L, Foulongne V, et al. The SARS-CoV-2 B.1.351 lineage (VOC β) is outgrowing the B.1.1.7 lineage (VOC α) in some French regions in April 2021. Eurosurveillance. 2021;26: 2100447. doi:10.2807/1560-7917.ES.2021.26.23.2100447

48. Benhamou W, Choquet R, Gandon S. The interplay between migration and selection on the dynamics of pathogen variants. J R Soc Interface. 2026;23: 20250867. doi:10.1098/rsif.2025.0867

49. Nationwide Commercial Laboratory Seroprevalence Survey | Data | Centers for Disease Control and Prevention. [cited 19 Aug 2025]. Available: https://data.cdc.gov/Laboratory-Surveillance/Nationwide-Commercial-Laboratory-Seroprevalence-Su/d2tw-32xv/about_data

50. COVID-19 Vaccinations in the United States, Jurisdiction | Data | Centers for Disease Control and Prevention. [cited 19 Aug 2025]. Available: https://data.cdc.gov/Vaccinations/COVID-19-Vaccinations-in-the-United-States-Jurisdi/unsk-b7fc/about_data

51. Taube JC, Susswein Z, Colizza V, Bansal S. Characterising non-household contact patterns relevant to respiratory transmission in the USA: analysis of a cross-sectional survey. Lancet Digit Health. 2025;7. doi:10.1016/j.landig.2025.100888

52. Blanquart F, Hozé N, Cowling BJ, Débarre F, Cauchemez S. Selection for infectivity profiles in slow and fast epidemics, and the rise of SARS-CoV-2 variants. Cooper BS, Davenport MP, editors. eLife. 2022;11: e75791. doi:10.7554/eLife.75791

53. Park SW, Bolker BM, Funk S, Metcalf CJE, Weitz JS, Grenfell BT, et al. The importance of the generation interval in investigating dynamics and control of new SARS-CoV-2 variants. J R Soc Interface. 2022;19: 20220173. doi:10.1098/rsif.2022.0173

54. Bureau UC. 2016-2020 5-Year ACS Commuting Flows. In: Census.gov [Internet]. [cited 4 May 2026]. Available: https://www.census.gov/data/tables/2020/demo/metro-micro/commuting-flows-2020.html

55. Social Distancing Metrics | SafeGraph Docs. In: SafeGraph [Internet]. [cited 13 May 2026]. Available: https://docs.safegraph.com/docs/social-distancing-metrics

56. Volz E, Mishra S, Chand M, Barrett JC, Johnson R, Geidelberg L, et al. Assessing transmissibility of SARS-CoV-2 lineage B.1.1.7 in England. Nature. 2021;593: 266–269. doi:10.1038/s41586-021-03470-x

57. Supasa P, Zhou D, Dejnirattisai W, Liu C, Mentzer AJ, Ginn HM, et al. Reduced neutralization of SARS-CoV-2 B.1.1.7 variant by convalescent and vaccine sera. Cell. 2021;184: 2201-2211.e7. doi:10.1016/j.cell.2021.02.033

58. Altarawneh HN, Chemaitelly H, Hasan MR, Ayoub HH, Qassim S, AlMukdad S, et al. Protection against the Omicron Variant from Previous SARS-CoV-2 Infection. N Engl J Med. 2022;386: 1288–1290. doi:10.1056/NEJMc2200133

59. Graham MS, Sudre CH, May A, Antonelli M, Murray B, Varsavsky T, et al. Changes in symptomatology, reinfection, and transmissibility associated with the SARS-CoV-2 variant B.1.1.7: an ecological study. Lancet Public Health. 2021;6: e335–e345. doi:10.1016/S2468-2667(21)00055-4

60. Di Domenico L, Sabbatini CE, Boëlle P-Y, Poletto C, Crépey P, Paireau J, et al. Adherence and sustainability of interventions informing optimal control against the COVID-19 pandemic. Commun Med. 2021;1: 1–13. doi:10.1038/s43856-021-00057-5

61. Weitz JS, Park SW, Eksin C, Dushoff J. Awareness-driven behavior changes can shift the shape of epidemics away from peaks and toward plateaus, shoulders, and oscillations. Proc Natl Acad Sci. 2020;117: 32764–32771. doi:10.1073/pnas.2009911117

62. Pangallo M, Aleta A, del Rio-Chanona RM, Pichler A, Martín-Corral D, Chinazzi M, et al. The unequal effects of the health–economy trade-off during the COVID-19 pandemic. Nat Hum Behav. 2024;8: 264–275. doi:10.1038/s41562-023-01747-x

63. Mastrandrea R, Fournet J, Barrat A. Contact Patterns in a High School: A Comparison between Data Collected Using Wearable Sensors, Contact Diaries and Friendship Surveys. PLOS ONE. 2015;10: e0136497. doi:10.1371/journal.pone.0136497

64. Salathé M, Kazandjieva M, Lee JW, Levis P, Feldman MW, Jones JH. A high-resolution human contact network for infectious disease transmission. Proc Natl Acad Sci. 2010;107: 22020–22025. doi:10.1073/pnas.1009094108

65. Génois M, Vestergaard CL, Fournet J, Panisson A, Bonmarin I, Barrat A. Data on face-to-face contacts in an office building suggest a low-cost vaccination strategy based on community linkers. Netw Sci. 2015;3: 326–347. doi:10.1017/nws.2015.10

66. Obadia T, Silhol R, Opatowski L, Temime L, Legrand J, Thiébaut ACM, et al. Detailed contact data and the dissemination of Staphylococcus aureus in hospitals. PLoS Comput Biol. 2015;11: e1004170. doi:10.1371/journal.pcbi.1004170

67. Vanhems P, Barrat A, Cattuto C, Pinton J-F, Khanafer N, Régis C, et al. Estimating Potential Infection Transmission Routes in Hospital Wards Using Wearable Proximity Sensors. PLOS ONE. 2013;8: e73970. doi:10.1371/journal.pone.0073970

68. Mossong J, Hens N, Jit M, Beutels P, Auranen K, Mikolajczyk R, et al. Social Contacts and Mixing Patterns Relevant to the Spread of Infectious Diseases. PLoS Med. 2008;5: e74. doi:10.1371/journal.pmed.0050074

69. Béraud G, Kazmercziak S, Beutels P, Levy-Bruhl D, Lenne X, Mielcarek N, et al. The French Connection: The First Large Population-Based Contact Survey in France Relevant for the Spread of Infectious Diseases. PLOS ONE. 2015;10: e0133203. doi:10.1371/journal.pone.0133203

70. Verelst F, Hermans L, Vercruysse S, Gimma A, Coletti P, Backer JA, et al. SOCRATES-CoMix: a platform for timely and open-source contact mixing data during and in between COVID-19 surges and interventions in over 20 European countries. BMC Med. 2021;19: 254. doi:10.1186/s12916-021-02133-y

71. Wong KLM, Gimma A, Coletti P, Paolotti D, Tizzani M, Cattuto C, et al. Social contact patterns during the COVID-19 pandemic in 21 European countries – evidence from a two-year study. BMC Infect Dis. 2023;23: 268. doi:10.1186/s12879-023-08214-y

72. Poletto C, Meloni S, Van Metre A, Colizza V, Moreno Y, Vespignani A. Characterising two-pathogen competition in spatially structured environments. Sci Rep. 2015;5: 7895. doi:10.1038/srep07895

73. Lion S, Gandon S. Spatial evolutionary epidemiology of spreading epidemics. Proc Biol Sci. 2016;283. doi:10.1098/rspb.2016.1170

74. Manica M, Litvinova M, Bellis AD, Guzzetta G, Mancuso P, Vicentini M, et al. Estimation of the incubation period and generation time of SARS-CoV-2 Alpha and Delta variants from contact tracing data. Epidemiol Infect. 2023;151: e5. doi:10.1017/S0950268822001947

75. Lion S, Gandon S. Evolution of class-structured populations in periodic environments. Evolution. 2022;76: 1674–1688. doi:10.1111/evo.14522

76. Figgins MD, Bedford T. Inferring variant-specific effective reproduction numbers from combined case and sequencing data. eLife. 2025;14. doi:10.7554/eLife.104802.1

77. Abousamra E, Figgins M, Bedford T. Fitness models provide accurate short-term forecasts of SARS-CoV-2 variant frequency. PLOS Comput Biol. 2024;20: e1012443. doi:10.1371/journal.pcbi.1012443

78. Britton T, Ball F, Trapman P. A mathematical model reveals the influence of population heterogeneity on herd immunity to SARS-CoV-2. Science. 2020;369: 846–849. doi:10.1126/science.abc6810

79. Gomes MGM, Ferreira MU, Corder RM, King JG, Souto-Maior C, Penha-Gonçalves C, et al. Individual variation in susceptibility or exposure to SARS-CoV-2 lowers the herd immunity threshold. J Theor Biol. 2022;540: 111063. doi:10.1016/j.jtbi.2022.111063

80. Loedy N, Wallinga J, Hens N, Torneri A. Repetition in social contacts: implications in modelling the transmission of respiratory infectious diseases in pre-pandemic and pandemic settings. Proc R Soc B Biol Sci. 2024;291: 20241296. doi:10.1098/rspb.2024.1296

81. Pung R, Firth JA, Russell TW, Rogers T, Lee VJ, Kucharski AJ. Temporal contact patterns and the implications for predicting superspreaders and planning of targeted outbreak control. J R Soc Interface. 2024;21: 20240358. doi:10.1098/rsif.2024.0358

82. Tkachenko AV, Maslov S, Elbanna A, Wong GN, Weiner ZJ, Goldenfeld N. Time-dependent heterogeneity leads to transient suppression of the COVID-19 epidemic, not herd immunity. Proc Natl Acad Sci. 2021;118: e2015972118. doi:10.1073/pnas.2015972118

83. Colizza V, Vespignani A. Epidemic modeling in metapopulation systems with heterogeneous coupling pattern: theory and simulations. J Theor Biol. 2008;251: 450–467. doi:10.1016/j.jtbi.2007.11.028

84. Kraemer MUG, Hill V, Ruis C, Dellicour S, Bajaj S, McCrone JT, et al. Spatiotemporal invasion dynamics of SARS-CoV-2 lineage B.1.1.7 emergence. Science. 2021;373: 889–895. doi:10.1126/science.abj0113

85. Tegally H, Wilkinson E, Tsui JL-H, Moir M, Martin D, Brito AF, et al. Dispersal patterns and influence of air travel during the global expansion of SARS-CoV-2 variants of concern. Cell. 2023;186: 3277-3290.e16. doi:10.1016/j.cell.2023.06.001

86. Kraemer MUG, Yang C-H, Gutierrez B, Wu C-H, Klein B, Pigott DM, et al. The effect of human mobility and control measures on the COVID-19 epidemic in China. Science. 2020;368: 493–497. doi:10.1126/science.abb4218

87. Wang J, Jiang L, Xu Y, He W, Zhang C, Bi F, et al. Epidemiology of influenza virus reinfection in Guangxi, China: a retrospective analysis of a nine-year influenza surveillance data: Characteristics of influenza virus reinfection. Int J Infect Dis. 2022;120: 135–141. doi:10.1016/j.ijid.2022.04.045

88. Manna A, Dall’Amico L, Tizzoni M, Karsai M, Perra N. Generalized contact matrices allow integrating socioeconomic variables into epidemic models. Sci Adv. 2024;10: eadk4606. doi:10.1126/sciadv.adk4606

89. Tizzoni M, Nsoesie EO, Gauvin L, Karsai M, Perra N, Bansal S. Addressing the socioeconomic divide in computational modeling for infectious diseases. Nat Commun. 2022;13: 2897. doi:10.1038/s41467-022-30688-8

90. Sajjadi S, Toranj Simin P, Shadmangohar M, Taraktas B, Bayram U, Ruiz-Blondet MV, et al. Structural inequalities exacerbate infection disparities. Sci Rep. 2025;15: 9082. doi:10.1038/s41598-025-91008-w

91. Jennings V, Lloyd-Smith B, Ironmonger D. Household size and the poisson distribution. J Aust Popul Assoc. 1999;16: 65–84. doi:10.1007/BF03029455

92. Eurostat. Average household size. In: Eurostat [Internet]. [cited 15 July 2025]. Available: https://ec.europa.eu/eurostat/databrowser/product/page/ILC_LVPH01

93. U.S. Census Bureau. Households and Families (Table S1101), 2023 American Community Survey 1-Year Estimates. [cited 15 July 2025]. Available: https://data.census.gov/table/ACSST1Y2023.S1101?q=household+size&g=010XX00US$0400000

94. Susswein Z, Valdano E, Brett T, Rohani P, Colizza V, Bansal S. Ignoring spatial heterogeneity in drivers of SARS-CoV-2 transmission in the US will impede sustained elimination. medRxiv. 2021; 2021.08.09.21261807. doi:10.1101/2021.08.09.21261807

95. Hastie T, Tibshirani R, Friedman J. The Elements of Statistical Learning. New York, NY: Springer; 2009. doi:10.1007/978-0-387-84858-7

96. Elbe S, Buckland-Merrett G. Data, disease and diplomacy: GISAID’s innovative contribution to global health. Glob Chall. 2017;1: 33–46. doi:10.1002/gch2.1018

97. Average size of U.S. households, by state 2021. In: Statista [Internet]. [cited 15 July 2025]. Available: https://www.statista.com/statistics/242265/average-size-of-us-households-by-state/

