## Supplementary text and figures. for "Contact structure and population immunity shape the selective advantage of emerging variants"

[†] Dr. Elisabeta Vergu has passed away during the completion of this work. Dr. Vergu has contributed to conceiving and designing the study, and developing the network-based multi-variant computational model.

#### Table of contents

**Estimate of the selection coefficient across US states** 3
**Additional results on the emerging variant simulations** 4
- Epidemic trajectories 4
- Association of the selection coefficient and the reproduction ratio 4
- Additional parameter exploration 6
- Variant absolute growth 6

**Emerging variant simulations: sensitivity analysis** 7
- Impact of initial seeding 7
- Impact of variation in the generation time of the emerging variant 8

Impact of mobility between populations 9
**Analysis of the US case study 12**
Correlation analysis stability over time 12
Effect of mobility coupling 13
**Consistency between model simulations and empirical estimates 14**
**References 16**

#### Estimate of the selection coefficient across US states

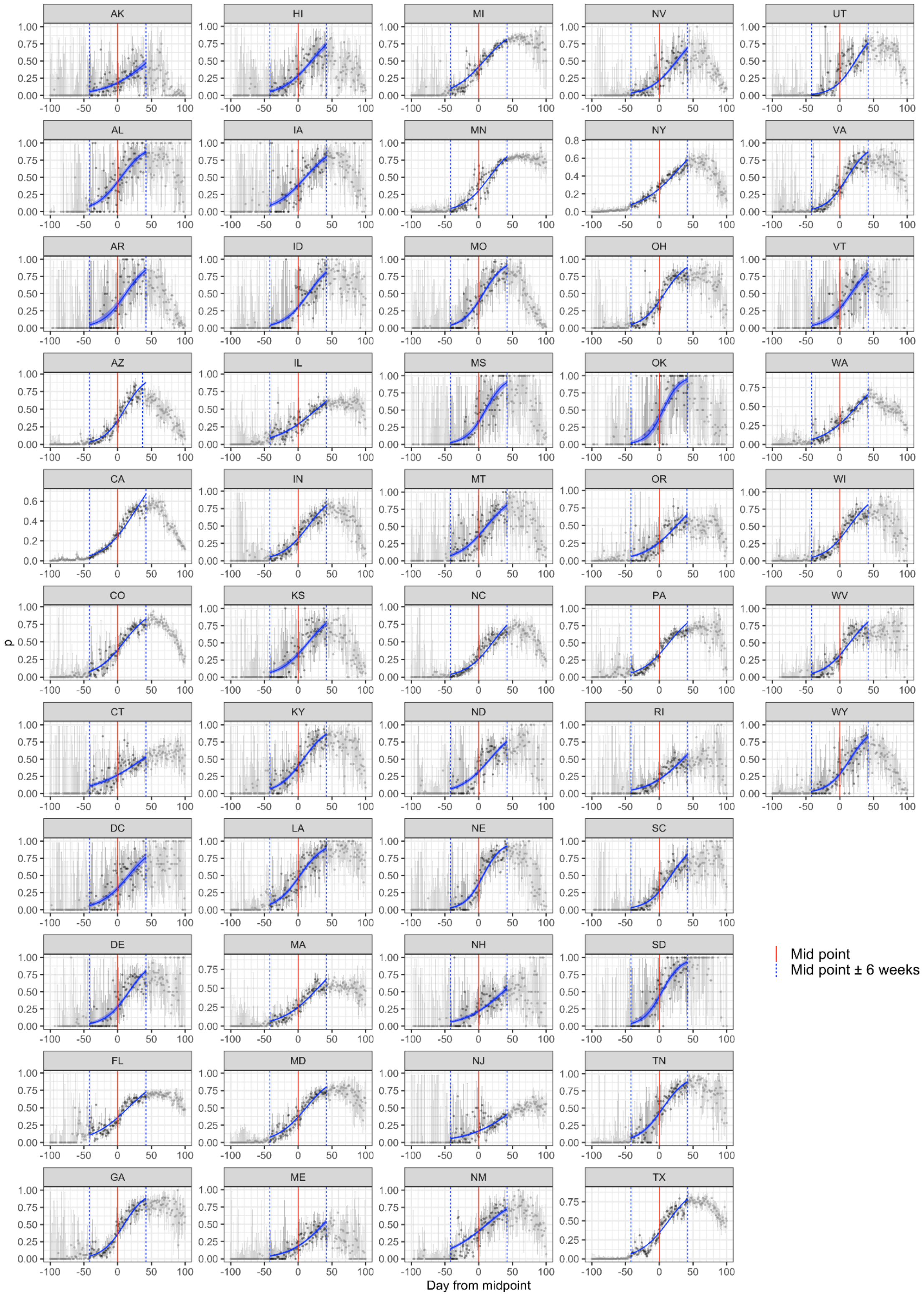


Fig S1. Frequency of the Alpha variant and the fitted logistic curve across US states. Observed Alpha variant frequencies across U.S. states are shown with their corresponding logistic fits, estimated using a 12-week time window. Error bars represent binomial confidence intervals for the estimated variant frequencies.

### Additional results on the emerging variant simulations

#### Epidemic trajectories

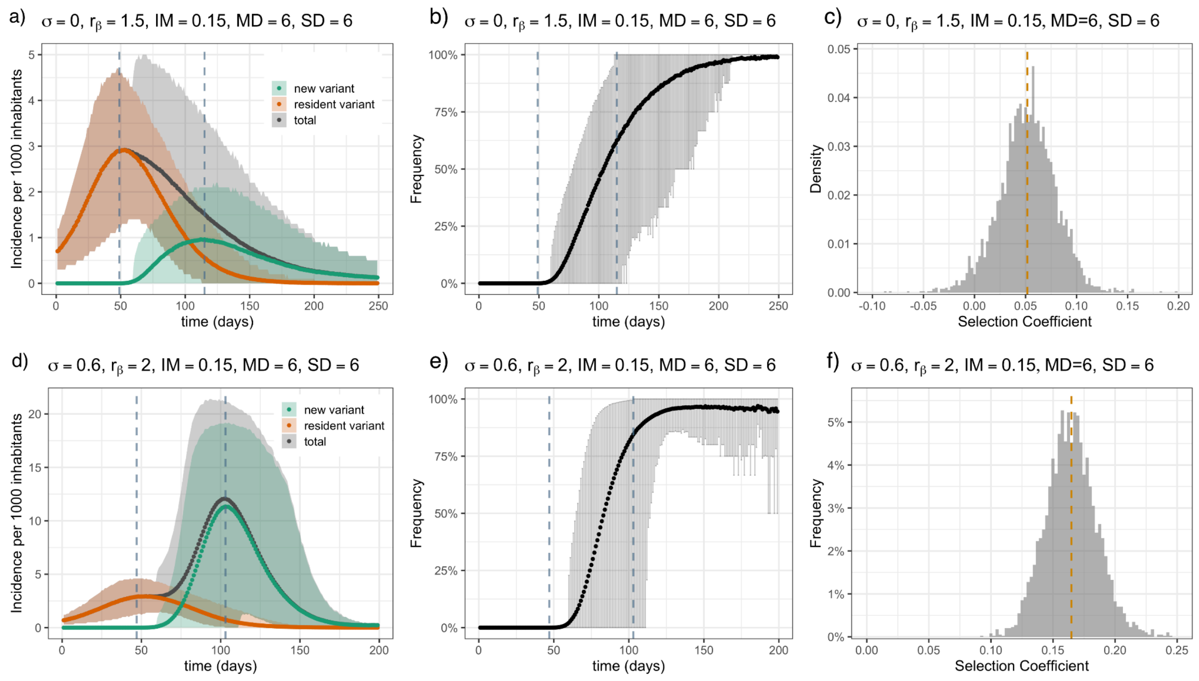


Fig S2. Examples of emergence dynamics. Panels (a, d) show the incidence of the resident variant, the emerging variant, and the total incidence as a function of time. Panels (b, e) show the frequency of the emerging variant, defined as the proportion of the incidence of the new variant to the total incidence. The shaded area shows the 95% confidence interval over 100 stochastic epidemic realisations for each of the 40 stochastic generations of the network with fixed parameters, for a total of 4000 runs. The vertical dashed lines represent the time window used to compute the selection coefficient. Panels (c, f) show the distribution of the computed selection coefficient of each realisation, representing the stochastic fluctuation of the estimated selection coefficient. The vertical orange line represents the mean value of the distribution. Panels (a, b, c) show a scenario with a mean selection coefficient of 0.052 (1/days), while panels (d, e, f) show a scenario with a mean selection coefficient of 0.165 (1/days). The key parameters of each scenario are indicated in the figure. Other parameters are: $\beta_r = 0.0175$, $\gamma = 0.143\ (1/7)\ days^{-1}$, size of the network $N = 10^4$.

#### Association of the selection coefficient and the reproduction ratio

In Fig. S3 we show that the selection coefficient is associated with the basic reproduction number of the contact network, computed as $R_0 = \frac{\beta}{\gamma}\left(\frac{SD_{ALL}^{2}}{MD_{ALL}} + MD_{ALL} - 1\right)$, particularly for sufficiently high values of immune evasion σ. In this regime, the relationship between the selection coefficient and $R_0$ becomes approximately linear and positive, with the selection coefficient increasing as $R_0$ increases.

This behavior can be understood from the structure of the susceptible network available to the emerging variant. For very low values of immune evasion $\sigma < 0.1$ and high levels of pre-existing immunity (IM), only a limited subset of individuals remains susceptible to the emerging variant. As a result, the effective susceptible network differs substantially from the original contact network, and the theoretical expression for $R_0$, derived for a fully susceptible population, becomes less predictive. The decrease with $R_0$ is due to the super blocker effect described in the main paper.

In contrast, for high immune evasion $\sigma$, a larger fraction of the population remains susceptible to the emerging variant. Consequently, the susceptible network more closely resembles the original fully susceptible contact network, making the theoretical prediction for $R_0$ more accurate and strengthening its association with the selection coefficient.

Additionally, when IM = 0, both variants emerge in a fully susceptible population. In this case, the susceptible network is identical for both variants, independent of the value of $\sigma$. Therefore, immune evasion does not affect the relationship between the selection coefficient and the reproduction number.

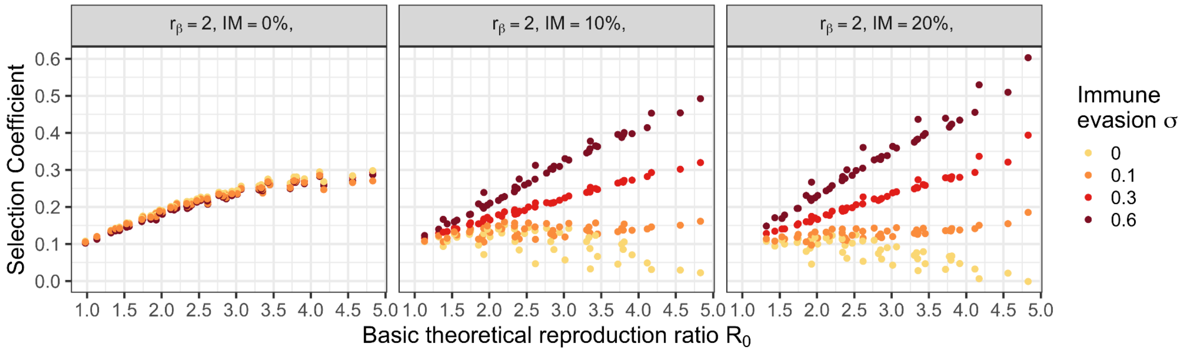


Fig S3. Association of the selection coefficient and the reproduction ratio. Selection coefficient (y-axis) as a function of basic reproduction ratio (x-axis) for different values of immunity (IM=0, 0.1, 0.2) with the transmissibility advantage of the emerging variant fixed at $r_\beta = 2$. Colors indicate the level of immune evasion (σ). Other parameters are: transmissibility of the resident variant $\beta = 0.0175$ and recovery rate $\gamma = 0.143\ (1/7)\ days^{-1}$.

#### Additional parameter exploration

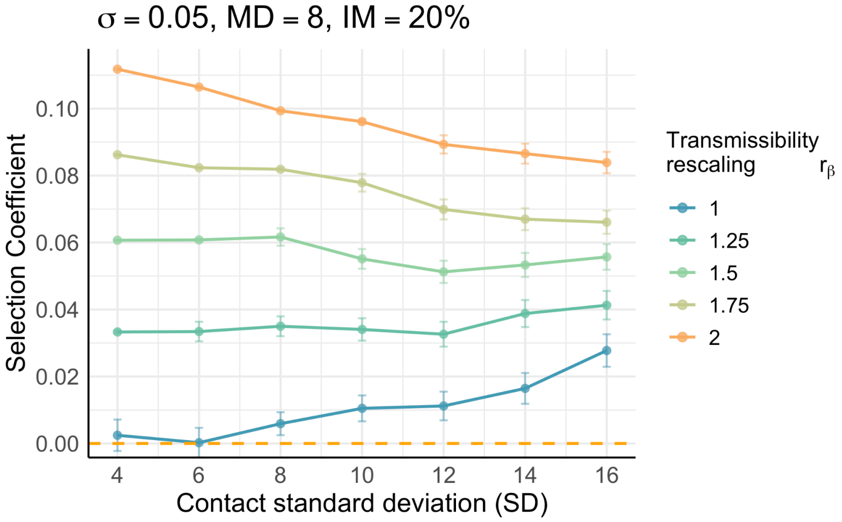


Fig S4. Selection coefficient (y-axis) as a function of standard deviation (SD) for a fixed immune evasion (σ = 0.05), mean of the number of contacts (MD = 8), and immunity level (IM=20%), where colours represent different transmissibility rescaling factors of the emerging variant. Points are means over 100 stochastic epidemic realisations for each of the 40 stochastic generations of the network with fixed parameters, for a total of 4000 runs.

#### Variant absolute growth

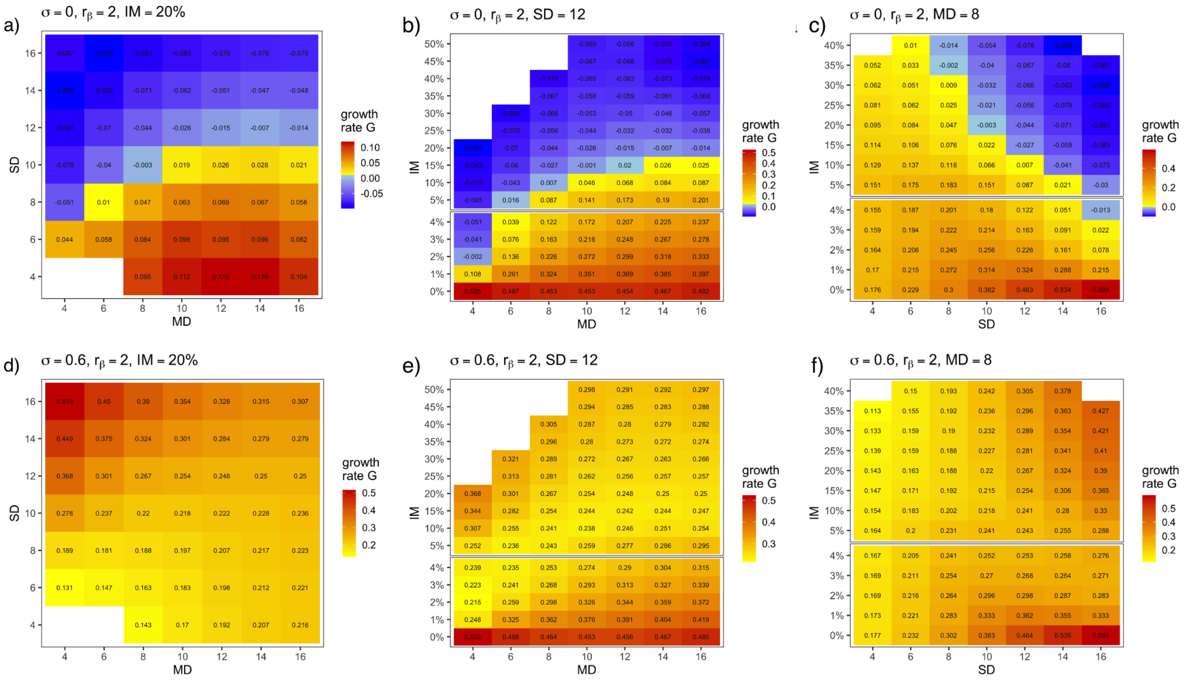


Fig S5. The heatmap of the growth rate under different parameter values. Panels (a, d) show results with fixed immunity (IM = 20%) for immune evasion σ = 0 (a) and σ = 0.6 (d). The x and y axes represent the contact mean (MD) and the contact standard deviation (SD), respectively. Panels (b, e) show results with fixed standard deviation (SD = 12) for σ = 0 (b) and σ = 0.6 (e). The x and y axes represent contact mean (MD) and immunity (IM), respectively. Panels (c, f) show results with fixed contact mean (MD = 8) for σ = 0 (c) and σ = 0.6 (f). The x and y axes correspond to SD and IM, respectively. Model parameters: transmissibility of the resident variant $\beta = 0.0175$, recovery rate $\gamma = 0.143\ (1/7)\ days^{-1}$, transmissibility advantage of the emerging variant $r_\beta = 2$, and network size N = $10^4$.

#### Emerging variant simulations: sensitivity analysis

##### Impact of initial seeding

We assessed the sensitivity of the results to the initial number of seeded infections. In the baseline parameterization, simulations were initialized with 50 randomly infected individuals for each variant. Here, we explored lower initial seeding levels of 5 and 10 individuals.

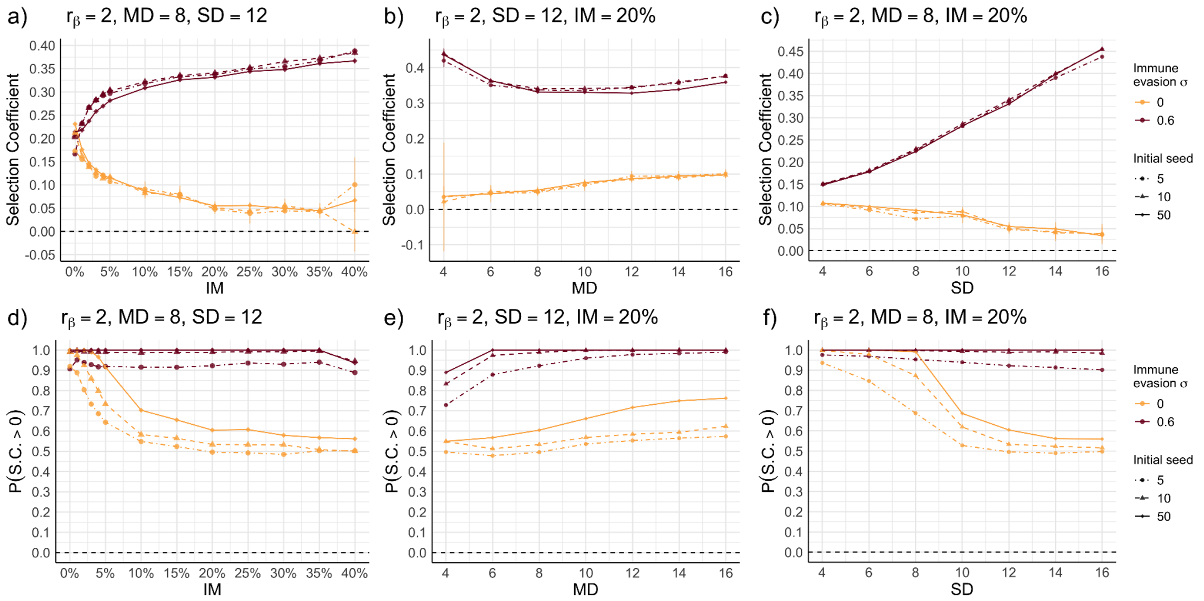


Fig S6. Impact of the initial number of seeders. Panel (a) shows the selection coefficient as a function of immunity (IM), for fixed values of the mean (MD = 8) and standard deviation (SD = 12) of the contact distribution. Panel (b) shows the selection coefficient as a function of the mean number of contacts (MD), for fixed values of immunity (IM = 20%) and standard deviation (SD = 12). Panel (c) shows the selection coefficient as a function of the standard deviation of contacts (SD), for fixed values of immunity (IM = 20%) and mean number of contacts (MD = 8). Panel (d) shows the probability of a positive selection coefficient as a function of immunity (IM), for fixed values of mean degree (MD = 8) and standard deviation of contacts (SD = 12). Panel (e) shows the same quantity as a function of MD, for fixed values of IM = 20% and SD = 12. Panel (f) shows the probability of a positive selection coefficient as a function of SD, for fixed values of IM = 20% and MD = 8. Colors indicate the level of immune evasion (σ), and marker shapes denote different assumptions for the initial number of seeded infections. Other parameters are: transmissibility of the resident variant $\beta = 0.0175$ and recovery rate $\gamma = 0.143\ (1/7)\ days^{-1}$.

##### Impact of variation in the generation time of the emerging variant

We explored the sensitivity of the results to variations in the generation time of the emerging variant. The generation time in the SIR model coincides with the recovery period. In the baseline parameterization, we assumed a recovery period of 7 days for both variants. Here, we also explored values of 6 and 8 days for the emerging variant.

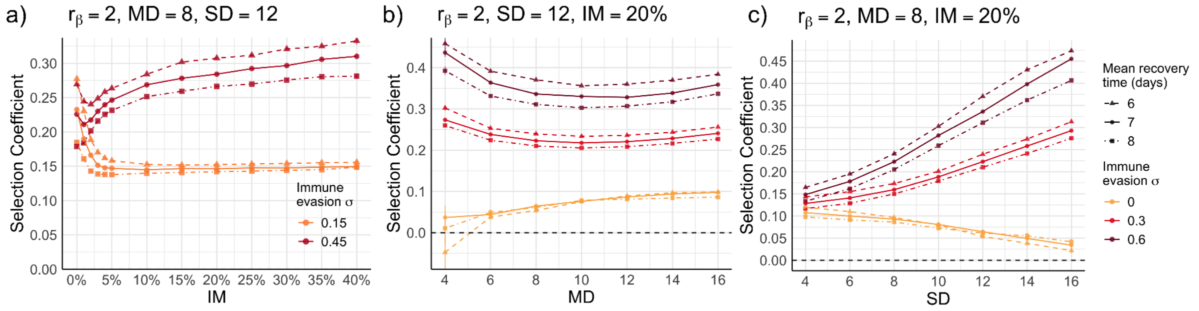


Fig S7. Impact of variation in the generation time (recovery time) of the emerging variant. Panel (a) shows the selection coefficient as a function of immunity (IM), for fixed values of the mean (MD = 8) and standard deviation (SD = 12) of the contact distribution. Panel (b) shows the selection coefficient as a function of the mean number of contacts (MD), for fixed values of immunity (IM = 20%) and standard deviation (SD = 12). Panel (c) shows the selection coefficient as a function of the standard deviation of contacts (SD), for fixed values of immunity (IM = 20%) and mean number of contacts (MD = 8). Colors indicate the level of immune evasion (σ), and marker shapes denote different assumptions for the mean recovery time of the emerging variant. Other parameters are: transmissibility of the resident variant $\beta = 0.0175$ and recovery rate $\gamma = 0.143\ (1/7)\ days^{-1}$.

**Impact of mobility between populations**

We explored how the dynamics of the emerging variant are affected by the continuous introduction of outside cases infected by either the emerging or the resident variant. To assess this effect, we developed a metapopulation model consisting of two populations, each represented as a contact network. Within each population $i$, susceptible individuals have a probability of becoming infected depending on the number of infected neighbours within $i$, like the baseline model. To this probability, we added an extra term, $\Lambda_i^j$, capturing the mobility coupling with $j$. The force of infection term, $\Lambda_i^j$, is proportional to the number of infectious individuals in the population $j$ and to a mobility scaling parameter that captures the fraction of individuals who travel. We modeled recurrent mobility fluxes, which are stronger and alter the local dynamics more strongly than long-range mobility. Approximating equations from (1, 2), $\Lambda_i^j$ accounts for (a) individuals from population $i$ traveling to population $j$, becoming infected there, and returning to $i$, and (b) infected individuals from population $j$ traveling to population $i$ and transmitting the infection. The mathematical expression of $\Lambda_i^j$ is

$$\Lambda_i^j = \beta\,\tau\,\left(\frac{MD_i}{N_i}\,p_t + \frac{MD_j}{N_j}\,p_t\right) I_j,$$

Where $MD_i$ and $MD_j$ are the mean degree of the network $i$ and $j$, respectively, $N_i = N_j = 10^4$ is the population of the two networks, $p_t$ is the traveling probability – we assumed for simplicity the traveling probability from $i$ to $j$ and from $j$ to $i$ equal – and $\tau$ represents the fraction of time individuals spend in the destination during a day. We set $\tau = 1/3\ days^{-1}$ as it is generally the case for commuting.

We modeled the emergence of a variant in the two networks and its co-circulation with the resident variant. While the intrinsic traits of the two variants were identical, some characteristics of the two populations could differ. In particular, the contact mean (MD), contact standard deviation (SD), and immunity at the time of emergence (IM) could vary between the two networks. We designated one population as the focal population and analyzed how the selection coefficient depends on its properties (MD, SD, and IM). We then assessed the extent to which this relationship is perturbed by incoming infections from the second population, characterized by parameters $MD_2$, $SD_2$, and $IM_2$.

Fig. S8 shows that the trends of the selection coefficient in varying IM are robust for any value of immune escape $\sigma$. Instead, for SD and MD, and only for the case of no immune escape, the trends vary in certain cases. In particular, the trends become non-monotonic for traveling probability $p_t = 0.01$ or higher. When MD and SD are greater than $MD_2$ and $SD_2$, however, the second network does not affect the selection coefficient of the focal network. In the figure, we consider the case in which the variant emerges in the second network first ($IM_2$<IM when IM is kept fixed). This case is more susceptible to yielding a perturbation of the focal network dynamics. Exploring the value $IM_2$ did not produce any appreciable effect.

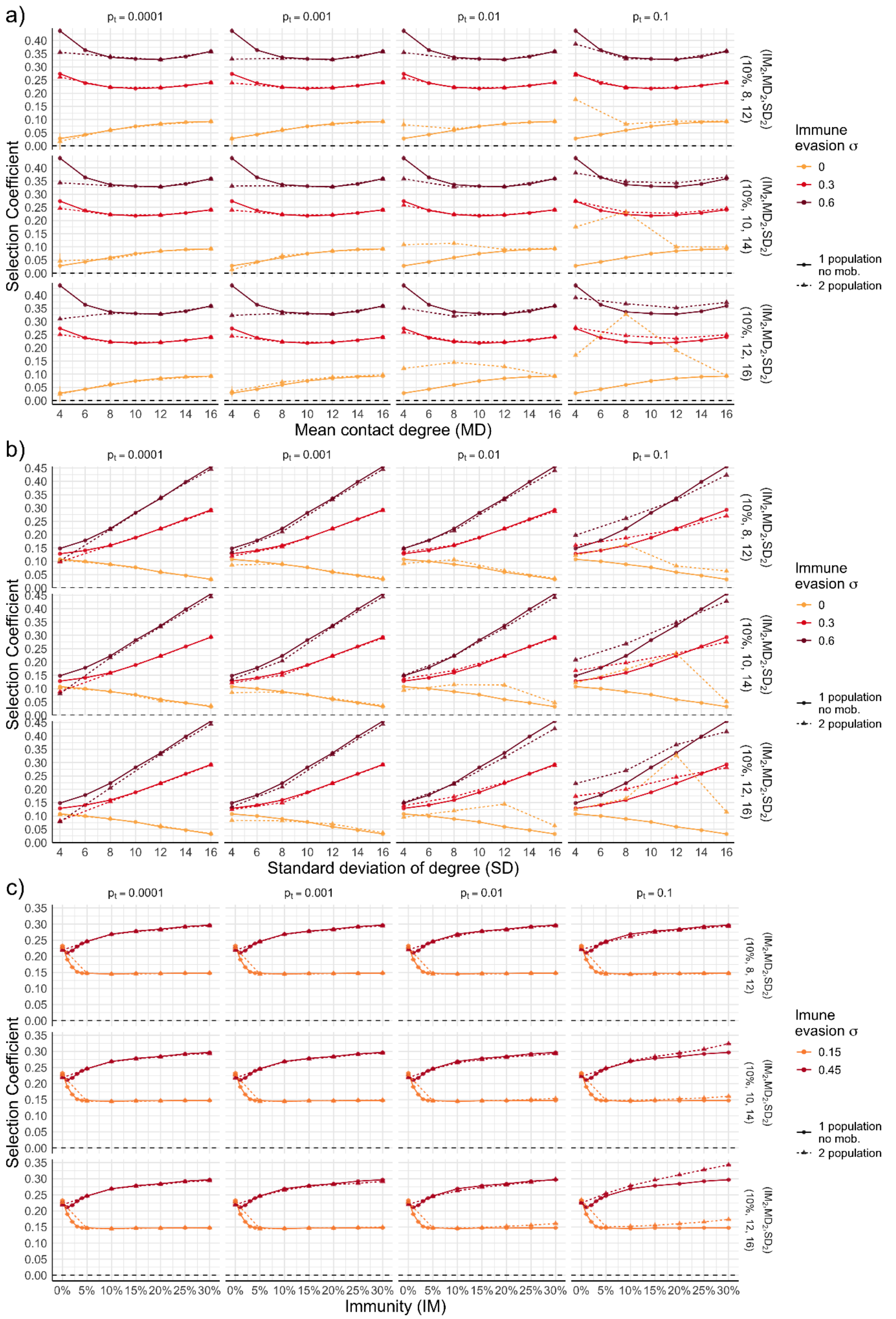


Fig S8. Impact of inter-population mobility on the selection coefficient. Each panel consists of a 4 × 3

grid of subplots. In panel (a), the selection coefficient is shown as a function of MD for fixed values of SD=12 and IM=20%. In panel (b), it is shown as a function of SD for fixed values of MD=8 and IM=20%. In panel (c), it is shown as a function of IM for fixed values of MD=8 and SD=12. Columns correspond to different traveling probabilities between the two patches (from left to right: 0.0001, 0.001, 0.01, and 0.1). Rows correspond to different parameter sets for the non-focal population: the first row corresponds to $IM_2$ = 10%, $MD_2$ = 8, and $SD_2$ = 12; the second row to $IM_2$ = 10%, $MD_2$ = 10, and $SD_2$ = 14; and the third row to $IM_2$ = 10%, $MD_2$ = 12, and $SD_2$ = 16. Solid lines represent the baseline scenario with a single population, while dashed lines correspond to a two-population system coupled through mobility.

#### Analysis of the US case study

##### Correlation analysis stability over time

We assess the sensitivity of the empirical results concerning the correlation between the selection coefficient of the Alpha variant and the characteristics of the population, namely MD, SD, and IM, across US states. In particular, we examine the robustness of these correlations with respect to the time window used to estimate the selection coefficient and covariates (IM, MD, and SD).

To this end, we shift the midpoint of the fitting time window from 21 days before to 21 days after the baseline midpoint. We considered a 4-week window centered on each midpoint and computed for each state the Alpha selection coefficient, the covariates, and the Pearson correlation between the selection coefficient and the covariates.

Fig. S9 shows how these correlations vary as the midpoint is shifted. The results were noisier, as expected, given the smaller time window used for the fit. However, the correlations with MD and SD were overall stable – i.e., consistently positive and in most cases statistically significant. The correlation with IM was less robust and in some cases not substantially different from zero.

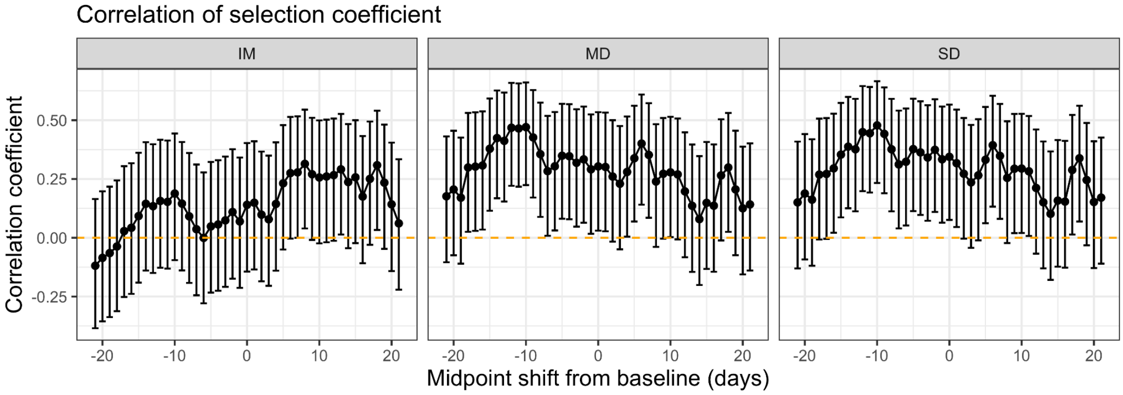


Fig S9. Time varying correlations. The figure shows the Pearson correlation between the selection coefficient and the covariates: contact mean (MD), contact standard deviation (SD), and immunity (IM). The y-axis represents the correlation coefficient, and the x-axis indicates the shift in the midpoint of the fitting window relative to the baseline (in days). The error bar represents 95% confidence interval in the calculation of the correlation coefficient.

**Effect of mobility coupling**

We quantified cross-state commuting fluxes as the probability of travel between pairs of states. The number of cross-state commuters between 2016 and 2020 was taken from (3). As the COVID-19 pandemic substantially altered mobility patterns, we rescaled this number by the reduction of mobility in workplaces provided in the Community Mobility Reports by Google (4). We computed commuting probabilities for the 2,269 state pairs with non-zero commuters by dividing the number of commuters by the population of the origin state. We found a median value of $2.59 \times 10^{-5}$ across all pairs. When we restricted it to neighbouring states, this increased approximately to $1.18 \times 10^{-3}$.

We also analyse a second mobility dataset containing all-purpose mobility at the county-day scale in the U.S. from January 4, 2021, to April 16, 2021. We aggregate these data to the state level across the study period to estimate the proportion of trips that occur between states (5). We found a median value of approximately $2.61 \times 10^{-4}$ across all pairs. When we restricted it to neighbouring states, this increased approximately to $5.87 \times 10^{-3}$.

According to both data sets, state pairs with high commuting probabilities often shared similar contact statistics, particularly among Northeastern states (e.g., Maryland, Massachusetts, Rhode Island, the District of Columbia, New Hampshire, Virginia, New York, and New Jersey). DC exhibited a disproportionately high cross-state mobility.

We performed robustness checks to verify that our correlation results were not biased by cross-state mobility coupling. We first recomputed Pearson correlation coefficients as in Fig. 4 of the main paper by removing DC. We found 0.6, $p=4.3 \cdot 10^{-6}$, for MD; 0.6, $p=3.8 \cdot 10^{-6}$, for SD; and 0.34, p=0.02, for IM–not substantially different, albeit slightly higher than baseline values (see main paper). We then removed states that were highly connected with states with substantially higher contact means. We used commuting data with a mobility threshold of $10^{-2}$, and a threshold for the difference in contact mean of 2. With this criterion, the removed states were Maryland, Minnesota, Washington, and DC. Pearson correlation values became 0.61, $p=4.7 \cdot 10^{-6}$, for MD; 0.61, $p=2.7 \cdot 10^{-6}$, for SD; and 0.33, p=0.03, for IM. We then removed 17 states that were highly connected (commuting with traveling probability $>10^{-2}$) with states with higher contact means (without any threshold). Pearson correlation values were in this case 0.60, p=0.0002, for MD; 0.60, p=0.0002, for SD; and 0.43, p=0.01, for IM. Finally, we repeated the same three tests as above using the all-purpose mobility dataset, and keeping the same threshold values for traveling probability and contact mean.

As mobility fluxes were higher with these data, the number of states that were removed from the analysis was higher. However, correlation values remained robust.

**Consistency between model simulations and empirical estimates**

We analyzed the correlation between the model simulations and the empirical estimates for the Alpha variant across US states. In particular, we ran simulations using the triplets of MD, SD, and IM values estimated for each state, together with the corresponding average household size, as inputs. We then computed the correlation between the simulated and empirical selection coefficients, as well as the correlations between the simulated selection coefficient and the covariates, mean number of contacts (MD), standard deviation of contacts (SD), and immunity level (IM).

We assumed a transmissibility advantage of $r_{\beta} = 1.5$, and an immune evasion level of $\sigma = 0.1$. The simulated and empirical selection coefficients across states were positively correlated (Pearson correlation = 0.55, $p < 10^{-4}$) after excluding Vermont, which was identified as an outlier (Fig. S10). Consistent with the empirical data, the simulated selection coefficient also exhibited positive correlations with all covariates: MD (Pearson correlation = 0.84, $p < 10^{-4}$), SD (Pearson correlation = 0.81, $p < 10^{-4}$), and IM (Pearson correlation = 0.40, $p < 0.02$).

All correlation values were slightly strengthened after excluding states with high commuting rates (Maryland, Minnesota, Washington, and the District of Columbia). In particular, the correlation between simulated and empirical selection coefficients increased to 0.57 ( $p = 4 \times 10^{-5}$), while the correlations between the simulated selection coefficient and MD, SD, and IM became 0.84 ($p < 10^{-13}$), 0.82 ($p < 10^{-11}$), and 0.44 ($p = 2.7 \times 10^{-3}$), respectively.

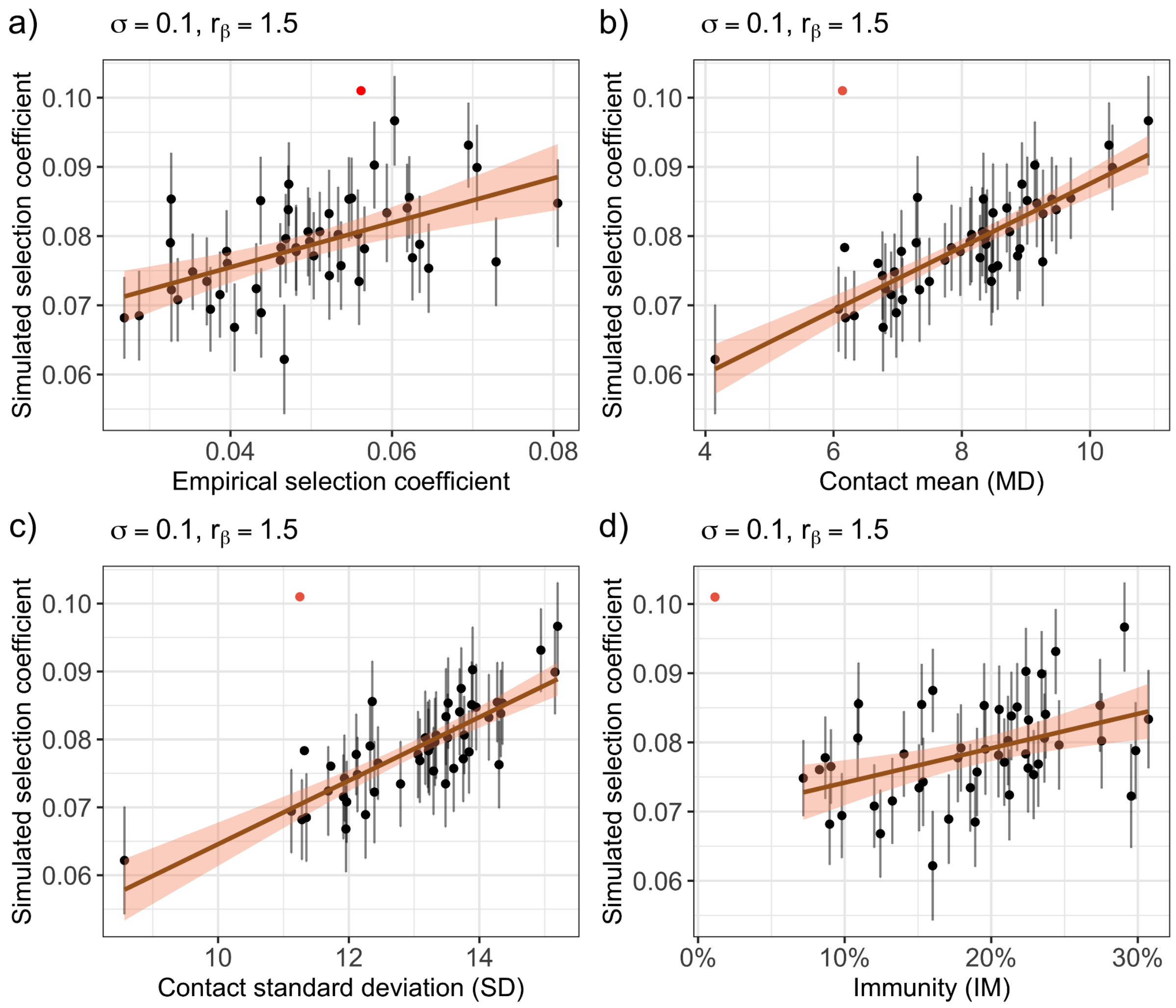


Fig S10. Model predictions of the selection coefficient across U.S. states. Input parameters correspond to the mean degree (MD), standard deviation (SD), and immunity level (IM) estimated for each state, together with the states' average household size. For each state, we constructed a synthetic contact network using these values to simulate emergence and computed the expected selection coefficient. Simulations are performed with a transmissibility advantage of the emerging variant ($r_\beta = 1.5$) and immune evasion (σ = 0.1). (a) Scatter plot of the simulated selection coefficient (y-axis) versus the empirical one across states (x-axis). (b) Simulated selection coefficients (y-axis) versus the mean number of contacts (MD, x-axis). (c) Simulated selection coefficients (y-axis) versus the standard deviation of contacts (SD, x-axis). (d) Simulated selection coefficients (y-axis) versus immunity levels (IM, x-axis). All correlations were computed after removing an outlier corresponding to the state of Vermont, which is indicated by a red dot in the scatter plots.

#### References


1. L. Sattenspiel, K. Dietz, A structured epidemic model incorporating geographic mobility among regions. *Math. Biosci.* **128**, 71–91 (1995).

2. M. J. Keeling, P. Rohani, Estimating spatial coupling in epidemiological systems: a mechanistic approach. *Ecol. Lett.* **5**, 20–29 (2002).

3. U. C. Bureau, 2016-2020 5-Year ACS Commuting Flows. *Census.gov*. Available at: https://www.census.gov/data/tables/2020/demo/metro-micro/commuting-flows-2020.html [Accessed 4 May 2026].

4. COVID-19 Community Mobility Report. *COVID-19 Community Mobil. Rep.* Available at: https://www.google.com/covid19/mobility?hl=en [Accessed 8 May 2026].

5. Social Distancing Metrics | SafeGraph Docs. *SafeGraph*. Available at: https://docs.safegraph.com/docs/social-distancing-metrics [Accessed 13 May 2026].
